# Multi-ancestry meta-analysis of keloids uncovers novel susceptibility loci in diverse populations

**DOI:** 10.1101/2025.01.28.25321288

**Authors:** Catherine A. Greene, Gabrielle Hampton, James Jaworski, Megan M. Shuey, Atlas Khan, Yuan Luo, Gail P. Jarvik, Bahram Namjou-Khales, Todd L. Edwards, Digna R. Velez Edwards, Jacklyn N. Hellwege

## Abstract

Keloids are raised scars that grow beyond original wound boundaries, resulting in pain and disfigurement. Reasons for keloid development are not well-understood, and current treatment options are limited. Keloids are more likely to occur in darker-skinned individuals of African and Asian descent than in Europeans. We performed a genome-wide association study (GWAS) examining keloid risk across and within continental ancestry groups, incorporating 7,837 cases and 1,593,009 controls. We detected 21 novel independent loci in the multi-ancestry analysis, including several previously associated with fibroproliferative disorders. Heritability estimates were 6%, 21%, and 34% for the European, East Asian, and African ancestry analyses, respectively. Genetically predicted gene expression and colocalization analyses identified 27 gene-tissue pairs, including nine in skin and fibroblasts. Pathway analysis implicated integrin signaling and upstream regulators involved in cancer, fibrosis, and sex hormone signaling. This investigation nearly quintuples the number of keloid-associated risk loci, illuminating biological processes in keloid pathology.

## INTRODUCTION

Keloids are raised scars that expand beyond the original wound boundaries and encroach on the surrounding skin.^1–3^ They are characterized by fibroproliferative derangement of the wound-healing process involving excessive deposition of collagen and overactive cell proliferation, though the precise etiology is not well-understood. The most common symptoms of keloids are pruritus (itchiness) and pain. More extensive or persistent keloids can cause disfigurement, and mobility issues may arise if keloids develop on joints. Notably, they have a tendency to recur after surgical excision and are frequently refractory to alternative treatments,^4^ necessitating an improved understanding of biological factors associated with susceptibility to excess scarring.

Keloids are most likely to affect darker-skinned individuals, particularly those of African or Asian descent. A sociodemographic study of keloids in the United States (US), with cases identified using structured and unstructured sources, documented a numerically higher proportion of Asian or Black patients with keloids.^5^ In the US, keloids occur in about 1 in 30 Black individuals, approximately a 20-fold increase in risk compared to White individuals.^6^ A recent study of excess scarring in the United Kingdom found prevalence estimates of 1.1%, 2.4%, and 0.4% for Asian, Black, and White patients, respectively.^7^ Other fibroproliferative diseases such as hypertension, sarcoidosis, and uterine fibroids display similar increased prevalence, and it has been suggested these conditions share biology with keloids.^8–10^

The limited genetic research conducted on keloids indicate they are a moderately heritable trait, as they can appear either sporadically or in people who have a family history. Family studies, however, have not identified a single genetic cause or mode of inheritance for keloids, fueling speculation that keloids are a complex trait with multiple susceptibility loci.^11^ Six genome-wide association studies (GWAS) since 2010, only one focused exclusively on keloids, have identified five distinct loci significantly associated with risk of keloids.^12–15^ However, these incorporated data from primarily European and East Asian ancestry populations; the earliest GWAS was a Japanese cohort study^12^ while the others were derived from analyses in the UK Biobank,^14,16,17^ FinnGen,^18^ and Biobank Japan.^13,15^ Only a recent analysis from the Million Veteran Program^19,20^ included individuals of African ancestry, who have the greatest burden of disease. Our group previously conducted a whole-exome association and admixture mapping study in a Black population,^21^ and we now present a GWAS meta-analysis of keloids mapping risk in diverse populations.

We aimed to capitalize on the sample diversity and data availability of Electronic Health Record (EHR)-linked biobanks by conducting multi-ancestry analyses in BioVU and eMERGE, in coordination with other large-scale data sources including the UK Biobank, FinnGen, Biobank Japan, and the US Veteran Administration’s Million Veteran Program (MVP). Because there are such striking health disparities in keloids susceptibility and severity, we conducted analyses stratified by ancestry group, depending on data availability, and present both multi-ancestry and ancestry-specific results. We additionally performed enrichment and gene expression analyses to investigate the functional consequences of disease-associated genes and gain novel insights into the biology of keloid scars.

## RESULTS

### GWAS Multi-ancestry Meta-analysis Results

We combined evidence of SNP-keloid associations through inverse variance-weighted fixed-effects meta-analyses, incorporating data from a total of 7,837 cases and 1,593,009 controls (**Figure 1**, **Table 1**) and limiting to common variants with minor allele frequency (MAF) ≥ 1%. We conducted meta-analyses both across and within ancestry groups, enabling the identification of keloids genetic risk factors exhibiting either multi-ancestry or ancestry-specific effects. The multi-ancestry analysis identified 26 significant (p ≤ 5×10^−8^), conditionally independent autosomal loci with support from multiple datasets, plus one additional locus of interest on the X chromosome (**Figure 2**, **Table 2**). Twenty-one of the 26 autosomal loci are novel genetic risk factors for keloids; we replicated four of five previously described loci and found additional support for a locus recently featured in a publication from the Million Veteran Program (**Supplementary Table 2**).^20^ Of the ten keloid-associated SNPs listed in the GWAS Catalog,^22^ seven were replicated at genome-wide significance in the multi-ancestry analysis.. Two SNPs had suggestive (p ≤ 1×10^−5^) evidence of association, and one SNP (rs1511412, *PRR23A*) lacked evidence of association outside the source Japanese population.

**Figure 1.**
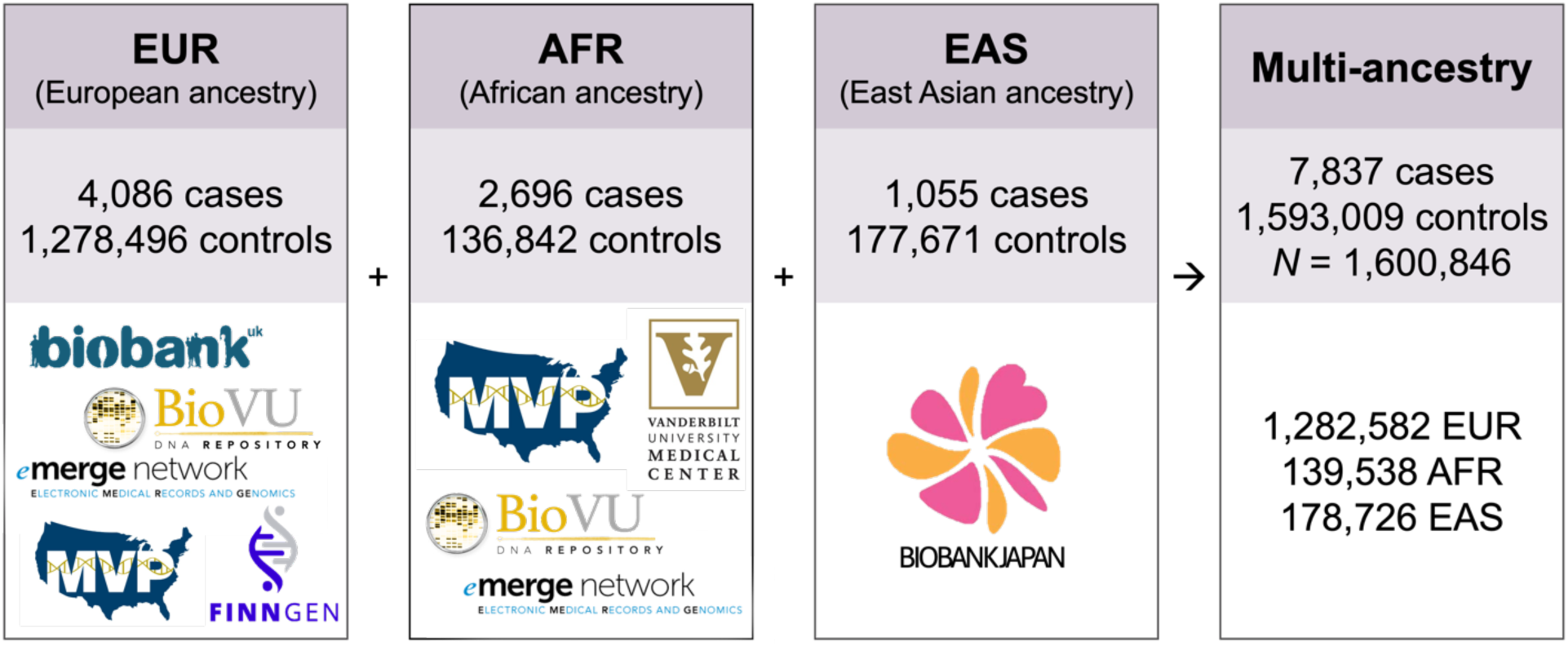
Summary of sample sizes and populations for meta-analyses.

**Figure 2.**
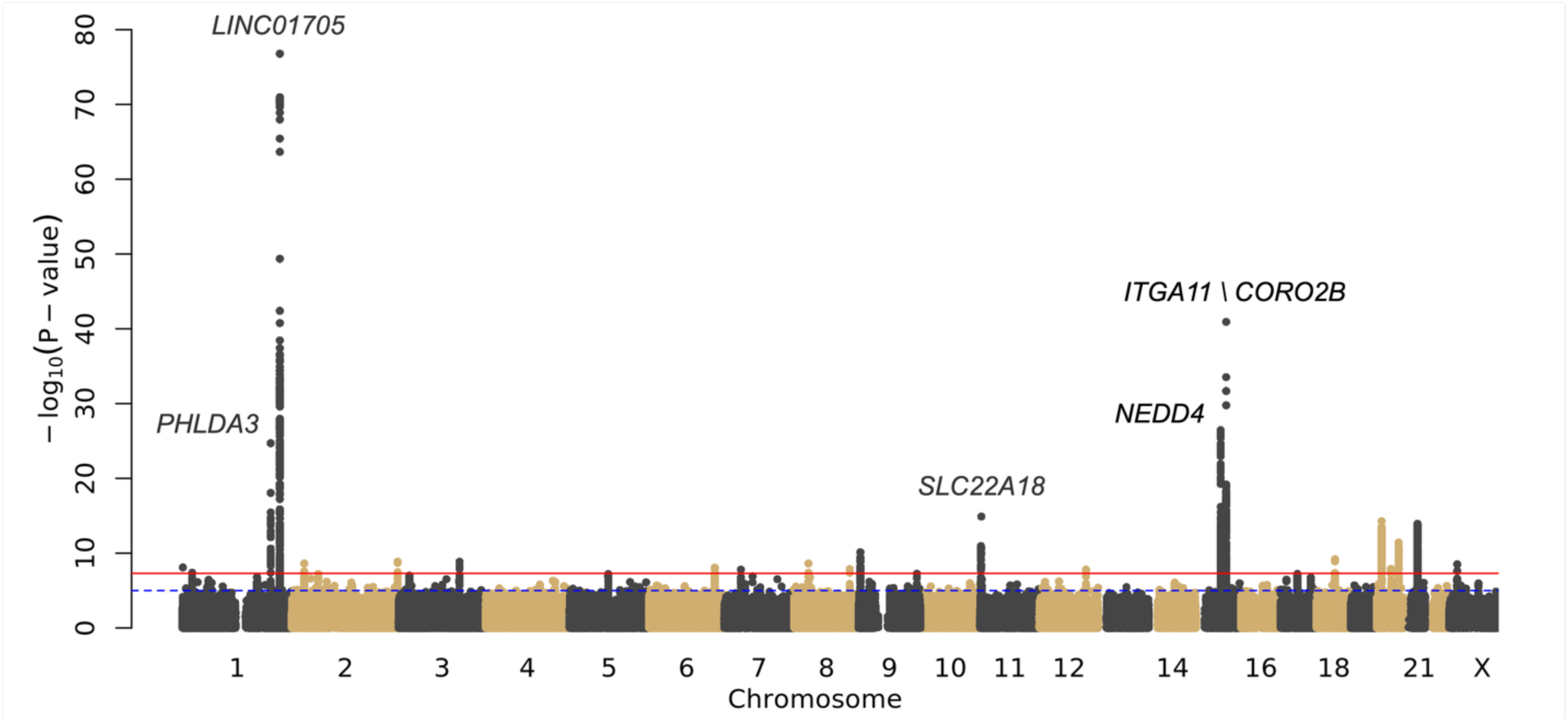
Manhattan plot showing the results of the keloids cross-ancestry meta-analysis. The top five most significant loci are labelled with the nearest gene. The red line signifies the traditional GWAS significance threshold of p< 5×10^-8^ while the blue dotted line signifies the suggestive threshold (p<1×10^-5^). Plot created using the fastman R library.

**Table 1.**
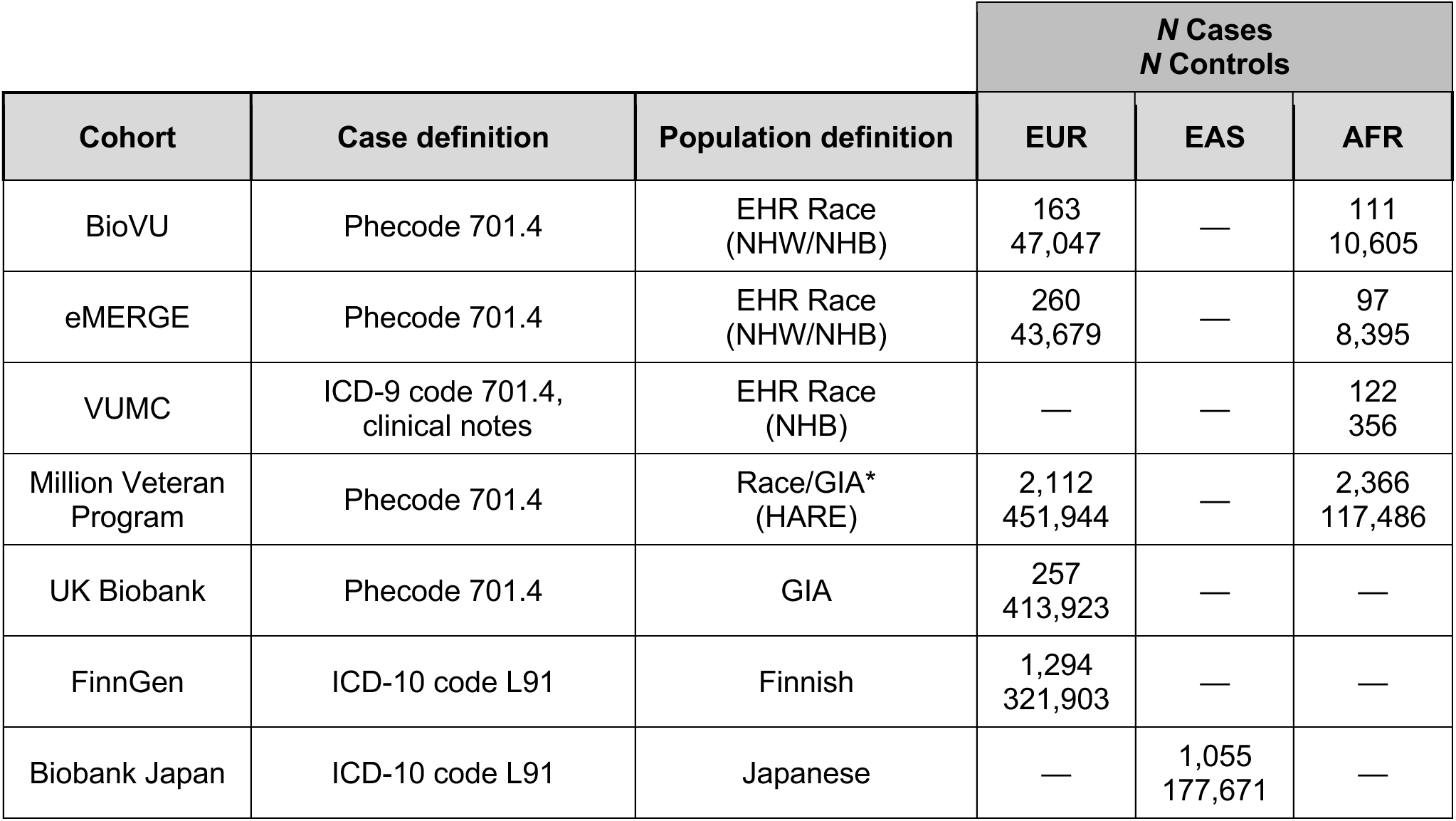
Cohort-specific sample sizes, case and population definitions. NHW=Non-Hispanic White; NHB=Non-Hispanic Black. *GIA=Genetically Inferred Ancestry.

**Table 2.** Independent lead SNPs in the multi-ancestry analysis, aligned to reflect increasing risk of developing keloids. Nearest Gene reports the FUMA-mapped gene for the lead SNP, except for *LINC01705* (mapped gene= RP11-400N13.1) to match previous keloid GWAS. SNP=rsid; Chr=Chromosome; BP=Base Pair; Risk=Risk Allele; Ref=Reference Allele; OR (95% CI)= Odds Ratio (95% Confidence Interval), effect conferred by the risk allele; Freq=weighted average of risk allele frequencies across all datasets. *This SNP represented in only one dataset; the rest have multiple supporting datasets.

| SNP | Chr:BP | Nearest Gene | Effect | Ref | OR (95% CI) | Freq | P-value |
| --- | --- | --- | --- | --- | --- | --- | --- |
| rs12568930 | 1:22702231 | *RP11-415K20.1* | C | T | 1.12 (1.07 — 1.16) | 0.24 | 5.92×10<sup>-09</sup> |
| rs35383942 | 1:201437832 | *PHLDA3* | T | C | 1.50 (1.39 — 1.61) | 0.08 | 1.91×10<sup>-25</sup> |
| rs10863683 | 1:222251089 | *LINC01705* | C | G | 1.41 (1.36 — 1.46) | 0.31 | 1.28×10<sup>-79</sup> |
| rs140707031 | 1:222291773 | *LINC01705* | G | A | 1.58 (1.33 — 1.89) | 0.02 | 9.43×10<sup>-09</sup> |
| rs6726716 | 2:28381833 | *BRE* | A | G | 1.12 (1.08 — 1.17) | 0.78 | 8.18×10<sup>-09</sup> |
| rs12989123 | 2:241252201 | *AC124861.2* | C | T | 1.16 (1.10 — 1.21) | 0.74 | 6.79×10<sup>-10</sup> |
| rs75826502 | 3:138837253 | *MRPS22:BPESC1* | C | G | 1.99 (1.59 — 2.49) | 0.04 | 1.41×10<sup>-09</sup> |
| rs244755 | 5:88095785 | *MEF2C* | T | C | 1.10 (1.06 — 1.14) | 0.54 | 2.29×10<sup>-08</sup> |
| rs6906384 | 6:149664540 | *TAB2* | G | A | 1.11 (1.07 — 1.15) | 0.56 | 5.36×10<sup>-10</sup> |
| rs2242026 | 7:37940286 | *EPDR1* | T | C | 1.15 (1.09 — 1.20) | 0.13 | 7.74×10<sup>-09</sup> |
| rs2919386 | 8:32555685 | *NRG1* | A | C | 1.22 (1.14 — 1.30) | 0.17 | 6.25×10<sup>-10</sup> |
| rs921721 | 8:126534536 | *RP11-136O12.2* | T | A | 1.11 (1.07 — 1.15) | 0.57 | 1.93×10<sup>-08</sup> |
| rs6476838 | 9:4287190 | *GLIS3* | T | C | 1.24 (1.16 — 1.32) | 0.80 | 1.20×10<sup>-10</sup> |
| rs686722 | 11:1891722 | *LSP1* | C | T | 1.14 (1.10 — 1.18) | 0.63 | 1.00×10<sup>-11</sup> |
| rs76024540 | 11:2920108 | *SLC22A18* | T | C | 1.44 (1.32 — 1.58) | 0.11 | 2.72×10<sup>-18</sup> |
| rs7297246 | 12:106119041 | *CASC18* | A | G | 1.16 (1.10 — 1.22) | 0.82 | 4.48×10<sup>-09</sup> |
| rs11632096 | 15:56210499 | *NEDD4* | G | A | 1.21 (1.17 — 1.26) | 0.32 | 1.11×10<sup>-27</sup> |
| rs34647667 | 15:68789866 | *ITGA11* | T | G | 1.39 (1.32 — 1.45) | 0.76 | 1.86×10<sup>-44</sup> |
| rs77685836 | 18:42241036 | *RP11-456K23.1* | C | G | 1.29 (1.19 — 1.40) | 0.93 | 2.07×10<sup>-09</sup> |
| rs2423510 | 20:10670079 | *RP11-103J8.1* | T | A | 1.12 (1.08 — 1.16) | 0.54 | 5.74×10<sup>-09</sup> |
| rs140716753 | 20:11106003 | *C20orf187* | A | C | 1.68 (1.46 — 1.95) | 0.03 | 2.39×10<sup>-14</sup> |
| rs4239705 | 20:11242516 | *RP4-734C18.1* | A | G | 1.15 (1.11 — 1.20) | 0.55 | 5.49×10<sup>-15</sup> |
| rs1205312 | 20:32849416 | *ASIP:RP4-785G19.5* | A | G | 1.28 (1.17 — 1.39) | 0.08 | 4.25×10<sup>-10</sup> |
| rs6091310 | 20:49984404 | *AL079339.1* | T | G | 1.13 (1.09 — 1.16) | 0.53 | 1.62×10<sup>-11</sup> |
| rs13051336 | 21:29816067 | *AF131217.1* | A | G | 1.16 (1.11 — 1.20) | 0.48 | 1.60×10<sup>-14</sup> |
| rs2832056 | 21:30132507 | *RNU6-872P* | G | T | 1.12 (1.08 — 1.17) | 0.52 | 9.34×10<sup>-10</sup> |
| rs769545468* | X:20810164 | *RP11-274G22.1* | A | G | 3.68 (2.39 — 5.65) | 0.01 | 3.06×10<sup>-09</sup> |

The most significant variant in the multi-ancestry analysis was rs10863683-C (p=1.52×10^-79^, Odds Ratio [OR] = 1.40 [95% Confidence Interval [CI] 1.35 — 1.45]), an intergenic variant near *LINC01705* (**Table 2**). This variant was originally detected in an East Asian population^13^ but was also significant in each of our ancestry-specific analyses. It was the most significant variant in the European ancestry analysis (p=1.32×10^-44^, OR = 1.39 [95% CI 1.32 — 1.45]) and the most significant variant at this locus in the African ancestry analysis (p=5.81×10^-19^, OR = 1.34 [95% CI 1.26 — 1.43]) (**Supplementary Table 4, Supplementary Figure 3**). A nearby intronic variant, rs11293015-G, was the most significant result in the East Asian ancestry analysis (p=1.39×10^-24^, OR = 0.60 [95% CI 0.54 — 0.66]) and may represent a conditionally independent locus in the Japanese population despite being in high Linkage Disequilibrium with rs10863683 in predominantly European populations (D’=0.26, r^2^=0.046 in Japanese; D’=0.969, r^2^=0.823 in European) (**Supplementary Table 4, Supplementary Figure 3**).^23^

The most significant novel locus in the multi-ancestry analysis was led by rs34647667-T (p=2.11×10^-44^, OR=1.39 [95% CI 1.33 — 1.46]), an intergenic variant located between *ITGA11* and *CORO2B* (**Table 2**). This SNP has the highest frequency in populations of African ancestry (gnomAD: MAF=0.35 in African/African American compared to 0.15 in European [non-Finnish] and 0.04 in East Asian [**Supplementary Table 4**])^23^ populations and was accordingly the most significant variant in the African ancestry-specific analysis (p=1.83×10^-32^, OR=1.48 [95% CI 1.39 — 1.58]). It also achieved significance in the European analysis (p=6.18×10^-14^, OR=1.30 [95% CI 1.21 — 1.38]), though the result at this locus was not significant in the East Asian analysis (rs34647667-T, p= 0.015) [**Figure 3, Supplementary Figure 6, Supplementary Table 4**].

**Figure 3.**
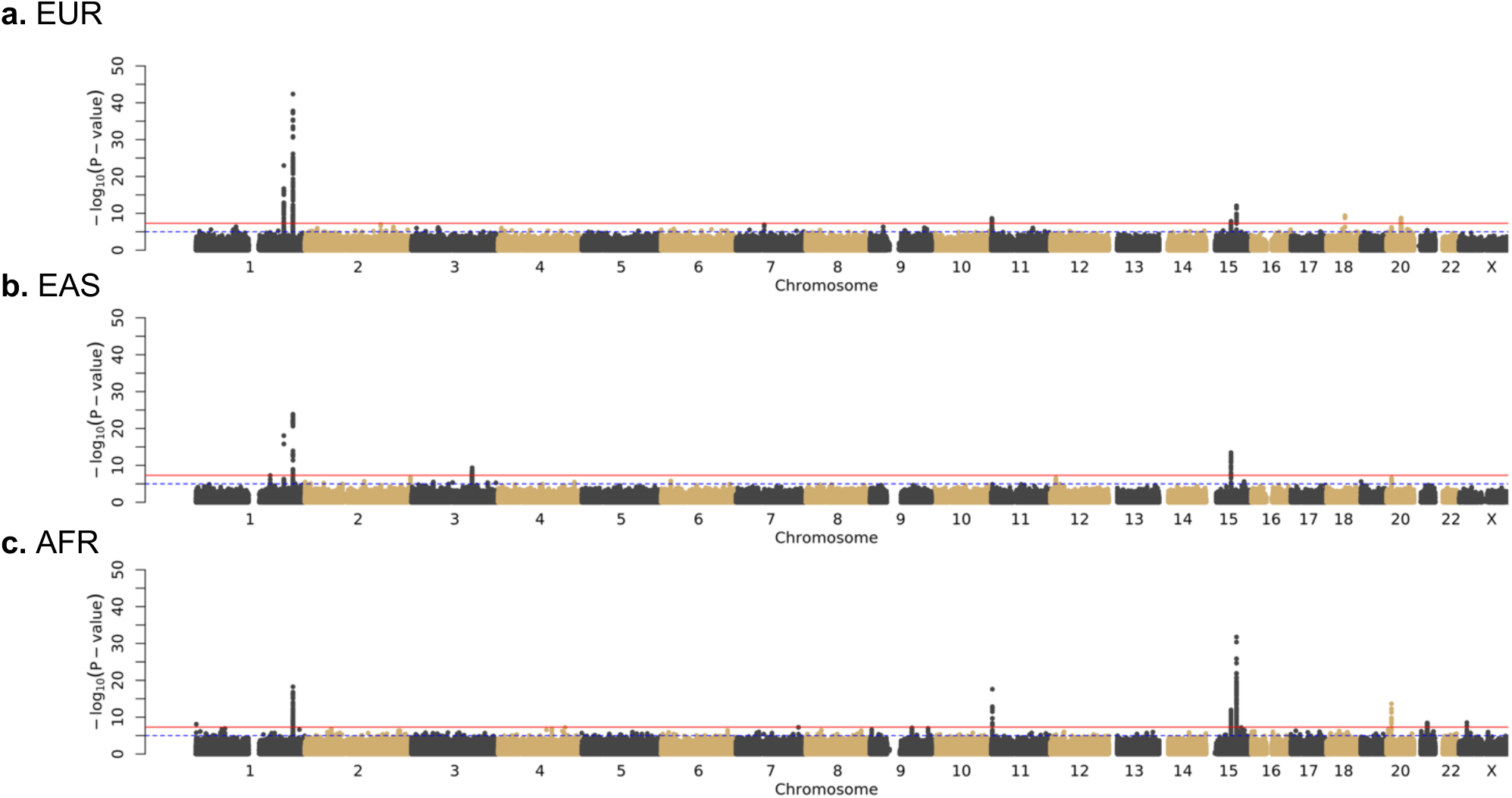
Manhattan plots showing the results of the keloids ancestry-stratified meta-analyses. **a.** European ancestry meta-analysis; **b.** East Asian ancestry meta-analysis; **c.** African ancestry meta-analysis. Plots created using the fastman R library.

### LDSC Results

We used Linkage Disequilibrium Score Regression (LDSC) to identify potential test statistic inflation and calculate the intercept for each analysis. Our multi-ancestry LDSC Intercept was 1.03, indicating that we do not have substantive confounding by population stratification (**Table 3, Supplementary Figure 1**). We also utilized LDSC to estimate SNP-based heritability of keloids from our ancestry-stratified GWAS summary statistics. Heritability was estimated to be approximately 0.06 in the European ancestry sample, 0.19 in East Asian ancestry, and 0.34 in African ancestry (**Figure 4**, **Table 3**). Heritability of keloids was highest in African ancestry, reflecting the observed pattern of disease prevalence in different populations.

**Figure 4.**
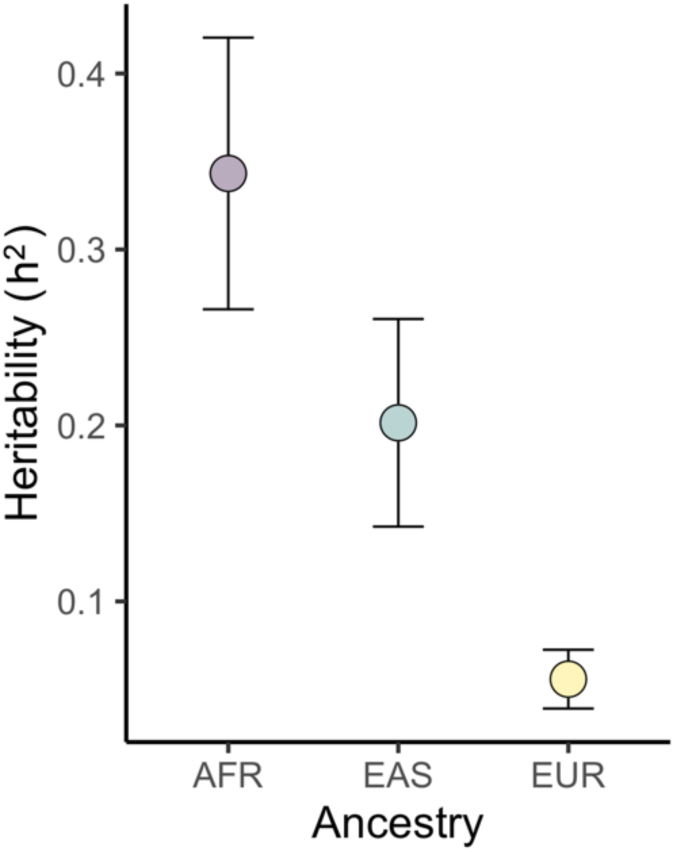
Heritability estimates.

**Table 3.** Intercept, λ_GC_, and SNP-based heritability (h^2^) values obtained with Linkage Disequilibrium Score Regression (LDSC). Summary statistics were filtered to approximately 1 million HapMap-defined SNPs in the pre-processing step, then merged to each respective LD reference to reach to final number of reference and regression SNPs listed in the last column.

| Ancestry | Intercept (SE) | $\lambda_{GC}$ | $h^2$ (SE) | *N* SNPs |
| --- | --- | --- | --- | --- |
| Multi-ancestry | 1.03 (0.006) | 1.07 | 0.06 (0.01) | 1,169,633 |
| EUR | 1.03 (0.007) | 1.05 | 0.06 (0.02) | 1,168,058 |
| EAS | 0.98 (0.007) | 1.00 | 0.19 (0.06) | 1,027,993 |
| AFR | 1.03 (0.007) | 1.07 | 0.34 (0.07) | 1,127,257 |

### Ancestry Comparison Results

Major results across ancestry-specific analyses were broadly consistent, though the relative strength of SNP-keloid associations tended to vary (**Figure 3**). For instance, the chromosome 1 consensus result described above, corresponding to the locus at *LINC01705*, was highly significant across all three ancestry-specific analyses but was superseded by other loci in the African and East Asian analyses (**Figure 3, Supplementary Figure 3, Supplementary Table 4**). A common result on chromosome 15 (rs11632096-A), mapping to *NEDD4,* was most significant in the East Asian (p=5.18×10^-13^, OR=0.71 [95% CI 0.65 — 0.78]) and African (p=3.14×10^-12^, OR=0.80 [95% CI 0.75 — 0.85]) analyses, but was less significant in the European analysis (p=7.07×10^-09^, OR=0.87 [95% CI 0.83 — 0.91]) (**Figure 3, Supplementary Figure 6, Supplementary Table 4**).

Many of the genetic risk loci for keloids also display heterogeneity among the ancestry-specific analyses. For example, the other strong result on chromosome 1, consisting of SNPs mapping to *PHLDA3*, is driven by the European (rs35383942-T, p=1.11×10^-23^, OR=1.49 [95% CI 1.38 — 1.61]) and East Asian analyses (rs192314256-C, p=8.74×10^-19^, OR=0.16 [95% CI 0.11—0.24]) (**Supplementary Figure 2**). There is no peak at this locus in the African ancestry analysis (rs35383942-T, p=0.005) though this discrepancy might be attributed to lower allele frequencies (MAF<1%) in African/African American populations.^23^ Further, the five distinct loci on chromosome 20 are driven by separate ancestry-specific analyses (**Table 2**). Three loci (rs2423510 [*JAG1*], rs6091310 [*NFATC2*], and rs4239705 [*RP4-734C18.1* / *AL049649.1*]) were only significant in the multi-ancestry analysis. The variant rs2423510 was not significant or even suggestive (p>1×10^-5^) in any of the ancestry-specific analyses; rs6091310 was only suggestive in the European analysis; and rs4239705 achieved suggestive significance in all three ancestry-specific analyses; but only the combined multi-ancestry evidence was sufficient to identify these keloid-associated loci. Two loci (rs140716753 [*C20orf187 / LINC02871*] and rs1205312 [*ASIP*]) were only significant in the African and European analyses, respectively, and may represent potential ancestry-specific genetic risk factors for keloids. Other ancestry-specific results include rs646315-T (*MRPS22*) on chromosome 3 in the East Asian analysis (p=4.63×10^-10^, OR=1.75 [95% CI 1.47 — 2.09]) (**Supplementary Figure 4**); rs76024540-T (*SLC22A18 / SLC22A18AS*) on chromosome 11 in the African analysis (p=2.54×10^-18^, OR=1.47 [95% CI 1.35 — 1.61]); and rs769545468-A on chromosome X in the African analysis (p=3.06×10^-9^, OR=3.68 [95% CI 2.39—5.65]).

### Consistency of Genetic Architecture across Ancestry Groups

To more broadly examine alleles influencing keloids risk across multiple ancestry-specific analyses, we compared both effect sizes and allele frequencies across European, East Asian and African populations for the lead SNPs identified in the multi-ancestry meta-analysis. The European and East Asian analyses had 19 lead SNPs in common; the European and African analyses had 22 lead SNPs in common; and the East Asian and African analyses had 19 lead SNPs in common (**Table 4**). Pairwise comparisons revealed moderate to strong correlations for both effect sizes (**Figure 5**) and allele frequencies (**Figure 6**), though notably this analysis could not include the five ancestry-exclusive alleles (**Tables 4 & 5**). We also performed the quantitative trait loci (QTL) sign test^24^ using the 26 lead autosomal SNPs to determine if allele frequency differences between populations constituted evidence of selection (**Supplementary Table 4**). No significant (p ≤ 0.05) differences were detected between the European and African analyses (p = 0.14), between the European and East Asian analyses (p = 0.14), or between the East Asian and African analyses (p = 0.12), suggesting that a greater number of keloid-associated alleles with frequency differences^25^ would be needed to reject the null hypothesis of no selection.

**Figure 5.**
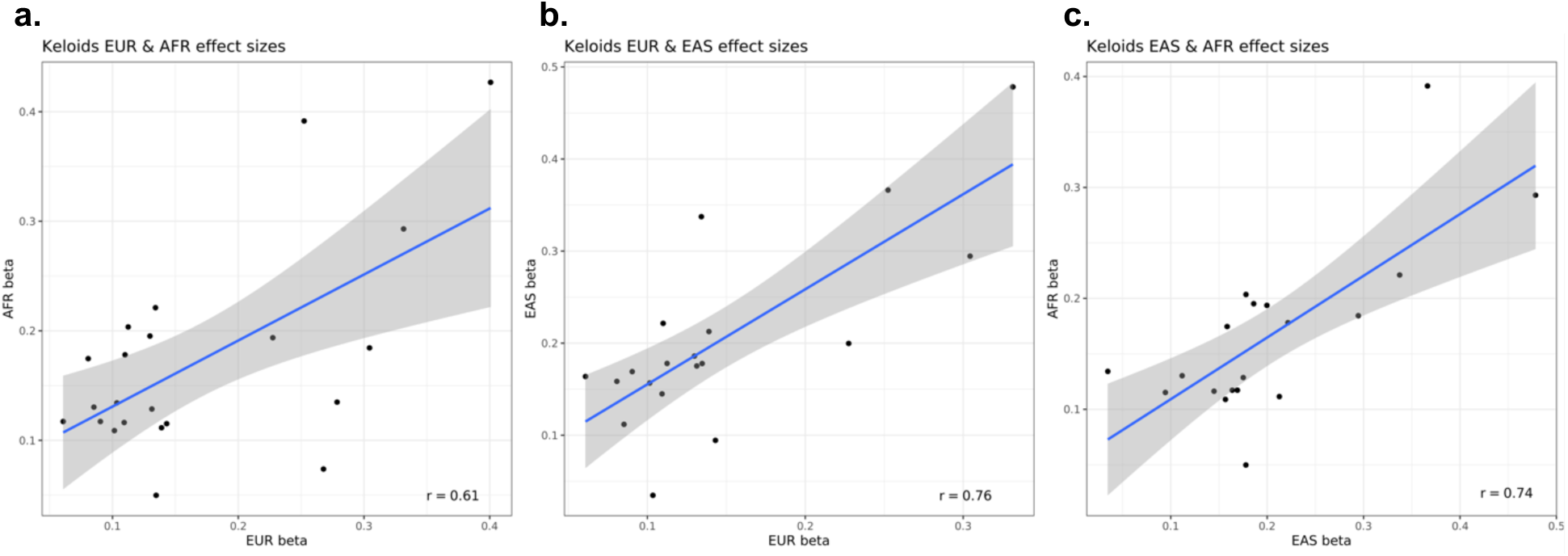
Effect size comparisons between ancestry-specific analyses, restricted to SNPs in common between each pairwise analysis. Alleles were aligned to the same risk direction for each comparison. r=Pearson correlation between SNP effect sizes; effect sizes represented as beta (β) values rather than as Odds Ratios, listed in Table 2.

**Figure 6.**
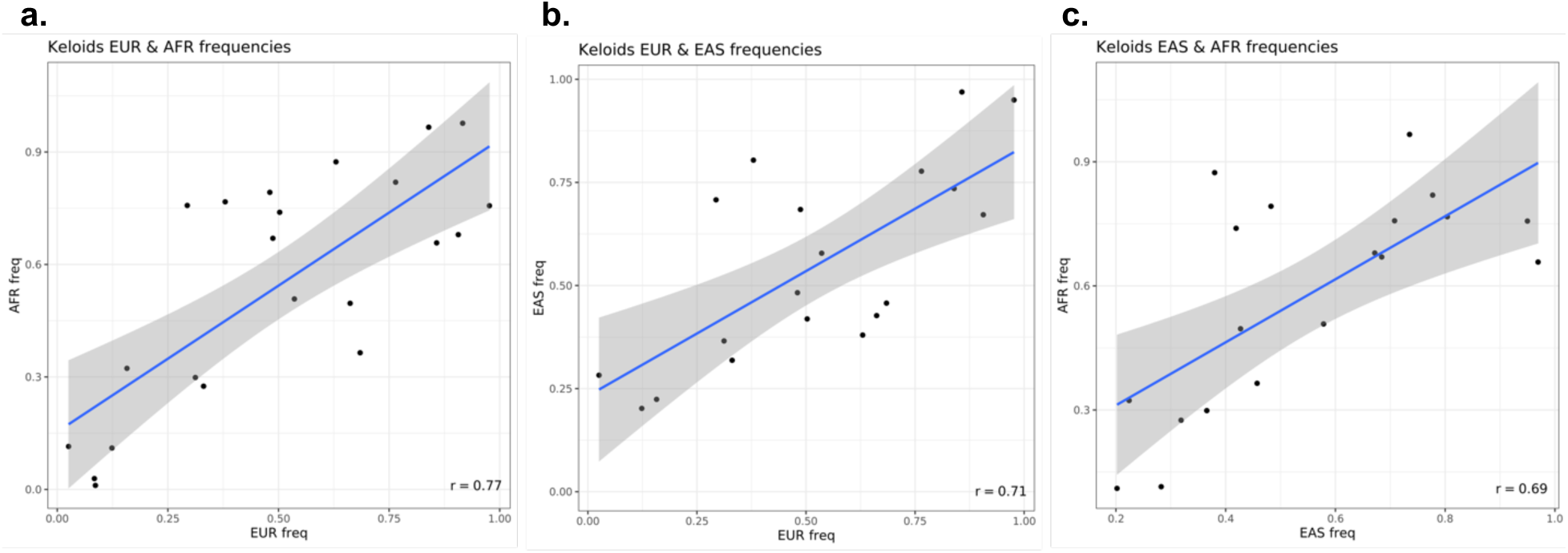
Allele frequency comparisons between ancestry-specific analyses, restricted to SNPs in common between each pair. Alleles were aligned to the same risk direction for each comparison. r=Pearson correlation between SNP allele frequencies; freq=allele frequency.

**Table 4.** Distribution of lead genome-wide significant SNPs across ancestry-specific analyses. Population-specific SNPs (ie, those represented at MAF>1% in only one ancestry-specific meta-analysis) are highlighted.

|  |  |  | Datasets represented |  |  |
| --- | --- | --- | --- | --- | --- |
| Chr | BP | SNP | EUR | EAS | AFR |
| 1 | 22702231 | rs12568930 | Y | Y | Y |
| 1 | 201437832 | rs35383942 | Y | N | Y |
| 1 | 222251089 | rs10863683 | Y | Y | Y |
| 1 | 222291773 | rs140707031 | N | N | Y |
| 2 | 28381833 | rs6726716 | Y | Y | Y |
| 2 | 241252201 | rs12989123 | Y | Y | Y |
| 3 | 138837253 | rs75826502 | N | Y | N |
| 5 | 88095785 | rs244755 | Y | Y | Y |
| 6 | 149664540 | rs6906384 | Y | Y | Y |
| 7 | 37940286 | rs2242026 | Y | Y | Y |
| 8 | 32555685 | rs2919386 | Y | Y | Y |
| 8 | 126534536 | rs921721 | Y | Y | Y |
| 9 | 4287190 | rs6476838 | Y | Y | Y |
| 11 | 1891722 | rs686722 | Y | Y | Y |
| 11 | 2920108 | rs76024540 | N | N | Y |
| 12 | 106119041 | rs7297246 | Y | Y | Y |
| 15 | 56210499 | rs11632096 | Y | Y | Y |
| 15 | 68789866 | rs34647667 | Y | Y | Y |
| 18 | 42241036 | rs77685836 | Y | N | Y |
| 20 | 10670079 | rs2423510 | Y | Y | Y |
| 20 | 11106003 | rs140716753 | N | N | Y |
| 20 | 11242516 | rs4239705 | Y | Y | Y |
| 20 | 32849416 | rs1205312 | Y | N | Y |
| 20 | 49984404 | rs6091310 | Y | Y | Y |
| 21 | 29816067 | rs13051336 | Y | Y | Y |
| 21 | 30132507 | rs2832056 | Y | Y | Y |
| X | 20810164 | rs769545468 | N | N | Y |

**Table 5.** Allele frequencies for population-specific SNPs. The highest population-level allele frequency for each SNP is shown, though each of these are the only populations in gnomAD where the frequency surpasses 1%. Risk=Risk Allele; Ref=Reference Allele; Freq=Allele Frequency.

| SNP | Effect | Ref | GWAS |  | gnomAD |  |
| --- | --- | --- | --- | --- | --- | --- |
|  |  |  | Population | Freq | Population | Freq |
| rs75826502 | C | G | EAS | 0.0430 | East Asian | 0.0113 |
| rs140707031 | G | A | AFR | 0.0237 | African/African American | 0.0177 |
| rs76024540 | T | C | AFR | 0.1082 | African/African American | 0.1188 |
| rs140716753 | A | C | AFR | 0.0332 | African/African American | 0.0356 |
| rs769545468 | A | G | AFR | 0.0105 | African/African American | 0.0124 |

### Functional Annotation of Keloid-Associated Genes

We utilized the online tool Functional Mapping and Annotation (FUMA) of GWAS^26^ to map variants to genes; functionally annotate variants; and perform gene set, tissue, and pathway enrichment analyses. Utilizing the SNP2GENE module, we found that the significant multi-ancestry GWAS results corresponded to 119 autosomal genes. Functional annotation of multi-ancestry significant variants revealed enrichment for SNPs in non-coding regions, consistent with the findings of most previous GWAS.^27^ Results were significantly enriched (E) for intronic noncoding (E=2.09, p=1.89×10^-134^) SNPs but were significantly depleted for intergenic (E=0.742, p=6.92×10^-65^) SNPs.

We also identified 12 nonsynonymous coding variants in LD (r^2^>0.1) with FUMA lead SNPs (Supplementary Table 14). Six SNPs achieved either suggestive or genome-wide significance and were mapped to *PHLDA3*, *TAB2/SUMO4*, *LSP1*, and *NEDD4*. Other nonsynonymous SNPs, though not significant in this study, map to genes previously associated with fibroproliferative diseases (e.g. *SFRP4*^28^) and may be of interest for future research.

Predicted effects of significant SNPs were estimated through Combined Annotation Dependent Depletion (CADD),^29^ RegulomeDB (RDB),^30^ and Chromatine 15 interaction values^31^ (E126, adult dermal fibroblasts) (**Supplementary Table 3**). In the multi-ancestry analysis, five of the 26 lead autosomal SNPs had CADD scores ≥ 12.37, considered the minimum value for pathogenic or highly deleterious SNPs.^32^ Three SNPs (rs35383942 [*PHLDA3*], rs34647667 [*ITGA11/CORO2B*], and rs11632096 [*NEDD4*]) had CADD scores>20, placing them in the top 1% of pathogenic SNPs. The maximum CADD value (24.3) was for rs35383942 (*PHLDA3*) which also was likely to affect binding (RDB = 2a) and located within an active transcription start site (E126 = 1). Variants rs34647667 (CADD = 21.5, RDB = 3a, E126 = 14) and rs11632096 (CADD = 20.1, RDB = 2b, E126 = 7) were similarly predicted to affect binding and have impacts on transcription.

We conducted gene set enrichment analyses using FUMA’s GENE2FUNC module, using the 119 mapped genes from the SNP2GENE module. The multi-ancestry tissue specificity analysis found that differentially expressed gene sets (GTEx v8 53 tissue types) characterizing the fallopian tubes (p=1.18×10^-4^),^33^ fibroblasts (p=3.61×10^-4^), and arterial tissues were significantly up-regulated (**Supplementary Table 6f**). Significant (False Discovery Rate (FDR) < 0.05) gene sets included: genes co-amplified with *MYCN* in primary neuroblastoma tumors and genes within amplicon 20q11 with copy number variations in breast tumors in both Curated Gene Sets and Chemical and Genetic Perturbation Gene Sets; and genes involved in *ERBB4* signaling events in Curated Gene Sets and All Canonical Pathways. Ancestry-specific pathway enrichment results were sparse but included the amplicon 20q11 gene set in the European analysis and genomically imprinted genes in the African ancestry analysis. Several significant multi-ancestry results in Gene Ontology (GO) cellular components and GO molecular functions related to muscle fibers and protein/lipid binding (**Supplementary Table 6a**). GWAS catalog reported traits had significant gene set enrichment for phenotypes including Dupuytren’s disease, calcium level, prothrombin time, breast and oral cavity cancers, and body mass index – though this list consisted of more skin- and tissue-related traits such as keloid and lobe attachment in the ancestry-specific analyses (**Supplement Table 6b-d**).

### Genetically Predicted Gene Expression (GPGE) Results

We next investigated the potential functional effects that keloid-associated variants may have on gene expression in various tissues using S-PrediXcan, in 49 tissues from GTEx v8.^34,35^ We detected 43 significant (p<1.7×10^-7^) gene-tissue pairs in the multi-ancestry analysis, corresponding to 20 unique genes across 25 tissues (**Supplementary Tables 7 & 8**). Results from three highly keloid-relevant tissues or cells (sun-exposed skin from the lower leg, not sun-exposed skin from the suprapubic region, and cultured fibroblasts) plus whole blood, included 11 significant (p<1.5×10^-6^) gene-tissue pairs, with some associated genes in multiple keloid-relevant tissues (**Figure 7**). The top result was for *LINC01705*; increased predicted expression of *LINC01705* in fibroblasts was associated with decreased risk of keloids (p=1.10×10^-20^, OR=0.65 [0.57 – 0.74]). Increased expression of both *PHLDA3* in sun-exposed skin (p=1.74×10^-14^, OR=2.27 [2.21 – 2.32]) and *NEDD4* in fibroblasts (p=5.90×10^-11^, OR = 1.91 [1.86 – 1.96]) was associated with increased risk of keloids. Increased predicted expression of the novel gene *LSP1* in fibroblasts (p=6.01×10^-8^, OR = 0.56 [0.55 – 0.58]) was associated with decreased risk of keloids. Although there was no significant GPGE result for *ITGA11*, the top novel result, we did observe a nearby association with *GLCE* in brain (amygdala, p=3.74×10^-19^, OR = 0.36 [0.35 – 0.37]) (**Supplementary Table 7**). We identified 27 gene-tissue pairs (nine in keloid-relevant tissues) with significant GPGE results and high posterior probability (PP>0.9) of colocalization.

**Figure 7.**
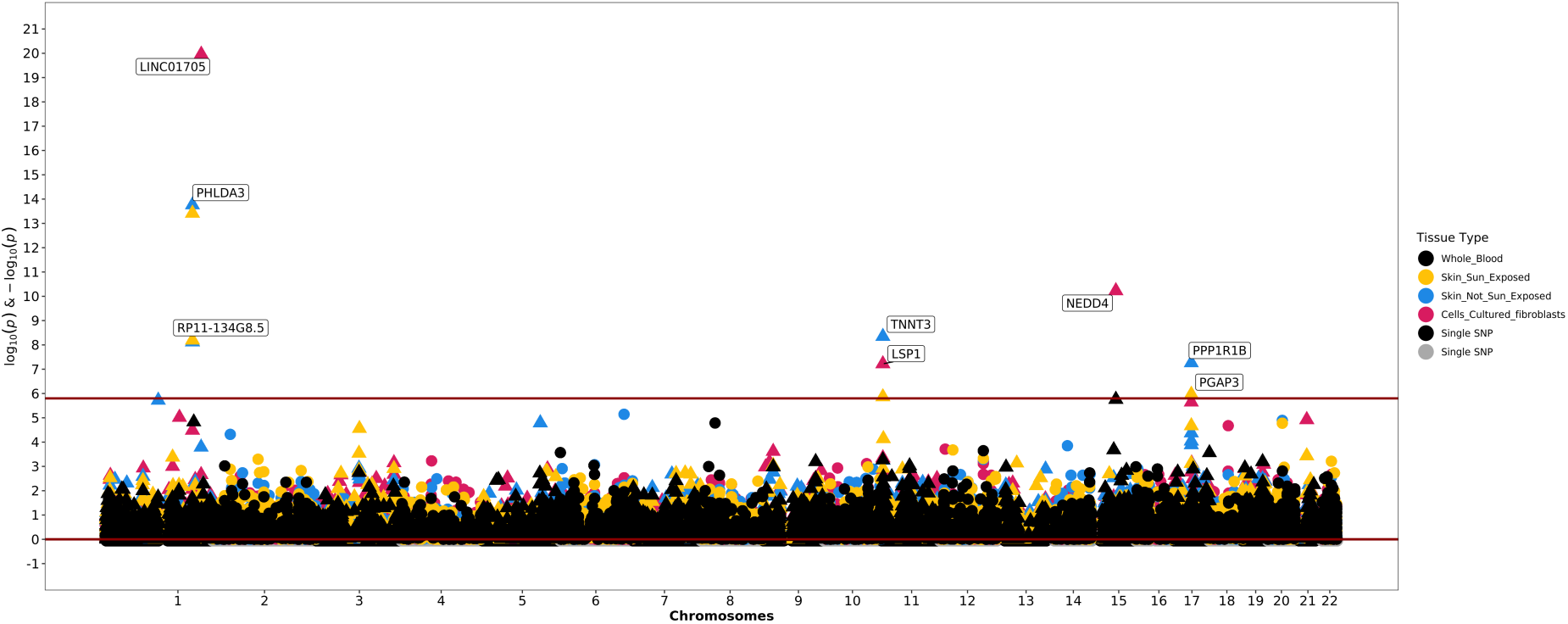
Cross-ancestry GPGE analysis, restricted to keloid-relevant tissues. Nine genes across four tissues are predicted to exhibit increased gene expression in response to keloids risk alleles. Significance threshold for the four keloid-relevant tissues = p<1.5×10^-6^.

The ancestry-specific GPGE analyses (all tissues) identified 41, eight, and nine significant gene-tissue pairs for the European, East Asian, and African analyses, respectively (**Supplementary Tables 7 & 8**). Only three genes were significant across all three ancestry-specific analyses: *LINC01705*, *NEDD4*, and *PRTG*. Other genes, notably *PHLDA3* and *RP11-134G8.5*, were significant in both the multi-ancestry and European analyses but not the East Asian and African analyses. Across the multi-ancestry and ancestry-specific analyses, we identified 22 unique (19 novel) genes with significant genetically predicted effects on gene expression. We used the union of the GWAS and GPGE gene lists as input for FUMA GENE2FUNC, which yielded significant (FDR<0.05) enrichment of genes involved in myogenesis, cardiac development, and cancer pathways (**Supplement Table 6e**).

### Ingenuity Pathway Analysis

We utilized QIAGEN’s Ingenuity Pathway Analysis software^36^ to determine potential upstream regulators and downstream biological functions of keloid-associated predicted gene expression, using nominally significant (p<0.05) GPGE results across the four meta-analyses. Twenty-five networks had significant enrichment of overlapping molecules in the multi-ancestry analysis, with top significant results including Carbohydrate Metabolism, Cell Cycle, Molecular Transport; Developmental Disorder, Gene Expression, Hereditary Disorder; and Cell Morphology, Cell-To-Cell Signaling and Interaction, RNA Post-Transcriptional Modification (**Supplementary Table 10**). The top enriched canonical pathways included Generic Transcription Pathway (-log[p] = 7.73) and Axonal Guidance Signaling (-log[p] = 7.27), consistent across all analyses (**Supplementary Table 11**). Other top pathways in the multi-ancestry analysis included Cardiac Hypertrophy Signaling (-log[p] = 5.64), Molecular Mechanisms of Cancer (-log[p] = 5.43), and Integrin Signaling (-log[p] = 4.88). The most significant upstream regulators, consisting of genes or other small molecules observed experimentally to affect expression of keloid-associated genes, included *HNF4A*, beta-estradiol, and dexamethasone across all analyses (**Supplementary Table 12**). Other top upstream regulators included *TP53* in the multi-, East Asian, and African ancestry analyses; *ESR1* in the multi-, European, and African ancestry analyses; and *TGFB1* in the European and East Asian ancestry analyses.

## DISCUSSION

We present a multi-ancestry meta-analysis of keloid scars incorporating large, diverse genetic datasets and notably includes populations with increased incidence of fibroproliferative disease. Both multi-ancestry and ancestry-stratified analyses were conducted to facilitate broad discovery of keloid-associated genomic risk loci and examine their biological impact through follow-up functional annotation, GPGE, and pathway enrichment analyses. Through our discovery meta-analysis, we replicated five known keloids risk loci and identified a further novel 21 loci. We additionally described 19 unique genes with genetically predicted effects on gene expression and evaluated genes’ roles in various biological processes and pathways that might contribute to disease.

We observed stark differences in estimated SNP-based heritability among the three ancestry-specific analyses, with keloids being most heritable in African ancestry populations (h^2^ = 0.34) and least heritable (h^2^ = 0.06) in European ancestry populations. These results reflect the observed pattern of keloids prevalence and suggest increased genetic susceptibility to keloids in some populations. This disparity in keloids genetic risk also provides some support for hypotheses suggesting their increased prevalence in African and Asian populations may be due to positive selection of fibroproliferative alleles,^8,9^ though we were unable to gather further supporting evidence through our comparisons of keloid effect allele frequencies via the QTL sign test.

In the multi-ancestry meta-analysis, we mapped keloid-associated variants to 119 genes, a substantial increase from the 12 total genes listed in the GWAS Catalog. We replicated associations at *LINC01705*, *PHLDA3*, *MRPS22/ BPESC1*, *SLC22A18/SLC22A18AS,* and *NEDD4* and identified numerous novel keloid-associated genes with diverse functions. Here we focus primarily on autosomal results, as BioVU alone included African ancestry X chromosome data representing the low frequency (MAF∼1% in African/African American populations, MAF<1% in other populations)^23^ lead variant rs769545468.

Many of the keloid-associated genes have functions related to tumor suppression. Although *LINC01705* has been proposed to be a regulatory factor underlying tumorigenesis, little is known about keloid-specific biological pathways or mechanisms despite this region being the most significant association in multiple studies.^12,13,15^ Overexpression of *LINC01705* was recently found to enhance cell migration and proliferation in breast cancer via regulation of *TPR*.^37^ It was also observed to be differentially expressed in colon cancer and positively correlated with two immunotherapy indicators, microsatellite instability and tumor mutational burden.^38^ The recently identified^20^ keloid locus *SLC22A18/SLC22A18AS* acts as a tumor suppressor in various cancers, including breast cancer and colorectal cancer.^39,40^ Novel genes associated with cancer phenotypes in the GWAS Catalog (accessed December 2024) include: *LSP1*^41,42^, *BRE / BABAM2*^43^, *ASIP*^44,45^, *TAB2*^41,42^, *NRG1*^46,47^, *TRIB1AL*^48^, *GLIS3*^49,50^, and *LINC02871*.^51^ Supporting these gene-level associations, an observational study examining risk of cancer development in patients with keloids found an overall increased cancer risk (OR = 1.49) in cases compared with controls, including increased risk for skin cancer (relative risk = 1.73).^52^

Other dermatologic and/or fibroproliferative conditions were associated with many of the novel genes, offering potential insights into keloid development. For instance, *LSP1* was previously shown to characterize a major fibroblast population regulating inflammation in normal human skin.^53^ Additionally, animal studies found that absence of *LSP1* promotes accelerated skin wound healing^54^ and alleviates asthmatic inflammation^55^ via reduced recruitment of inflammatory cell types. The region around *NEDD4* was previously found to contain an admixture mapping peak associated with keloid formation in African Americans, with the most significant result at *MYO1E*.^21^ The novel keloid-associated gene *ITGA11*, located roughly a megabase downstream of *NEDD4*, encodes a collagen receptor and is involved in regulation of the profibrotic TGFβ-signaling pathway.^56^ *ITGA11* was the top novel result and top result overall in the African ancestry analysis and was previously associated with various kinds of organ fibrosis,^56^ uterine fibroids^15,57,58^ and Dupuytren’s disease,^59–61^ an abnormal accumulation of fibrotic tissue resulting in contracture of the hand. Uterine fibroids are of particular interest due to their hypothesized shared etiology with keloids, supported by patterns in comorbidity among the two conditions.^9^ We did not observe a significant GPGE result at *ITGA11*, though the nearby associations at *GLCE* might indicate that effects may be attenuated due to shortcomings of the current gene expression model (see *Considerations*).

Other relevant GWAS catalog phenotypes for these keloid-associated genes include several skin cancers; skin and hair pigmentation traits; male pattern baldness; earlobe morphology; and rosacea. Previously associated fibroproliferative conditions include asthma; glaucoma; hypertension and other blood pressure traits; uterine fibroids; and Dupuytren’s disease. Other previously associated conditions consisted of musculoskeletal traits such as osteoporosis and decreased bone mineral density, which have been found in cross-sectional studies to co-occur with keloids.^7,62^

We observed several results that were consistent across pathway analyses, notably *TP53* and *TGFβ*. These genes have important roles in signal transduction; cell growth and differentiation; cell proliferation and migration; cell cycle signaling; and apoptotic pathways; and have both been robustly associated with fibrosis and cancers.^63,64^ Dexamethasone, a glucocorticoid that downregulates *VEGF* expression, is used to suppress angiogenic activity as a first-line treatment for keloids.^65,66^ *ESR1*, an estrogen receptor, is potentially reflective of the sex-specific effects of keloids, as cases of worsening keloids in pregnancy or after puberty in females have been documented.^67–69^ The most significant networks largely involved cellular signaling and cancer pathways, supporting pathway enrichment results obtained through the FUMA GENE2FUNC analyses.

### Considerations and Conclusions

There are several considerations that may affect interpretation of results. The most important of these is cohort composition, as population definitions varied among source datasets (**Table 1**). Some biobanks utilized methods to obtain genetically inferred ancestry, while others relied on stratification of biobank populations by EHR-reported race (i.e., Black and White) and ethnicity (non-Hispanic). Case/control definitions also varied slightly between datasets. Some definitions may have also resulted in the inclusion of hypertrophic scars, a condition of lesser severity that is difficult to clinically distinguish from keloids and which cannot be excluded with current code-based definitions. Though there are known differences in the pathophysiology of keloids and hypertrophic scars, it is unclear if their etiology is entirely separate or if they exist on a continuum of dysregulated wound healing. Differing sample sizes among ancestry groups, previously discussed in context of specific analyses, impact our ability to draw conclusions regarding common versus ancestry-specific impacts of genetic risk factors for keloids. Current gene expression models are also characterized by a deficiency of samples from populations disproportionately impacted by fibroproliferative disease. Future work will ideally incorporate more diverse samples or be enabled by studies directly examining keloid-affected tissue. Despite these considerations, we have described numerous novel genetic risk factors affecting excess scarring across different populations.

Through this large multi-ancestry meta-analysis of keloids risk, we identified several novel genomic risk loci contributing to keloid development. Some had evidence of impacts on gene expression in keloid-relevant tissues. Many of the genes identified have previously documented roles in fibrosis and wound healing, with dysregulation affecting cellular functions contributing to diverse disease phenotypes. Our findings also offer preliminary support for genetic risk factors that vary based on ancestry, with stark differences in heritability indicating heterogenous genetic susceptibility to keloid scarring depending on the ancestral population. An expanded understanding of the genetic architecture of keloids may assist in the identification of alternative treatments for scar management. This investigation may also serve to encourage future functional research examining mechanisms of keloids and highlight opportunities for studies of fibroproliferative disease.

## MATERIALS AND METHODS

### Study Populations

The total multi-ancestry meta-analysis amounted to approximately 1.6 million individuals across seven source biobanks. UK Biobank,^16^ FinnGen,^18^ Biobank Japan,^15^ and the Million Veteran Program (MVP)^19^ all were previously utilized in studies of keloids. The remainder, which included both BioVU and VUMC resources and the eMERGE Network were novel datasets for use in keloids genetic discovery. The MVP comprised a little over 85% of the samples in the African-ancestry meta-analysis.

All datasets utilized code-based definitions for the identification of keloid cases (**Table 1**). BioVU, eMERGE, MVP, and UK Biobank used phecode 701.4 to identify cases. VUMC used ICD-9 code 701.4 in addition to clinical notes to identify cases.^21^ FinnGen and Biobank Japan used ICD-10 code L91 to identify cases. The prevalence of keloids in each dataset is reflective of their relative rarity in most populations; 0.3% of the European ancestry samples, 0.6% of the East Asian ancestry samples, and 2% of the African ancestry samples were keloid cases.

### BioVU and eMERGE GWAS Analysis

BioVU is the DNA biorepository at Vanderbilt University Medical Center, linked to de-identified electronic health records in the Synthetic Derivative.^70^ DNA derived from peripheral blood samples was genotyped on a custom Illumina Multi-Ethnic Genotyping Array (MEGA-ex; Illumina Inc., San Diego, CA, USA) platform and were imputed through Trans-Omics for Precision Medicine (TOPMed).^71^ The Electronic Medical Records and Genomics (eMERGE) network is a national network supported by the National Human Genome Research Institute that connects data from EHR-linked DNA biorepositories across the country for large-scale collaborative research efforts promoting genomic medicine.^72–74^ eMERGE samples were genotyped using various platforms and were imputed through Haplotype Reference Consortium (HRC).^75^ For both BioVU and eMERGE, analyses were stratified according to EHR race. Quality control procedures were performed separately for non-Hispanic White and non-Hispanic Black individuals. Variants were limited to those with INFO scores>0.5, minor allele count>5, and above 95% genotyping rate. Chromosome X variants from BioVU were limited to those with INFO scores>0.7. The phecode 701.4 (keloids and hypertrophic scars) to identify cases in both BioVU and eMERGE (**Table 1**). Association analyses were performed with plink2^76^, adjusted by sex and the top 10 PCs.

### Multi-ancestry and Ancestry-specific Meta-analyses

We conducted fixed-effects inverse-weighted meta-analysis of keloids GWAS datasets using METAL.^77^ The multi-ancestry meta-analysis utilized all datasets (European, African, and East Asian ancestry) from each data source. The European ancestry meta-analysis utilized datasets with European or White population descriptors, including the UK Biobank and FinnGen, as well as results from the MVP, BioVU, and eMERGE. The African-ancestry meta-analysis utilized datasets with African or Black population descriptors, including results from the MVP, BioVU, and eMERGE. We performed GWAS meta-analyses of keloids including variants with MAF ≤ 1%, first conducted with all datasets for the cross-ancestry analysis, then also stratified by broad ancestry group. We used the traditional genome-wide significance threshold of 5.0×10^-8^ and set the suggestive threshold at 1.0×10^-5^.

### Ancestry Comparison

Variants were restricted to those achieving genome-wide significance in at least one analysis and were additionally limited to lead SNPs to avoid bias introduced by LD. Variants were aligned as needed using the EasyQC^78^ R package prior to conducting effect size and allele frequency comparisons between pairs of ancestry-specific analyses. Allele frequencies were acquired from gnomAD,^23^ using European (non-Finnish) values for EUR; East Asian values for EAS; and African/African American values for AFR. Pearson’s correlation was calculated for each pairwise comparison and are reported in **Figures 5 & 6**.

### FUMA

Summary statistics for the multi-ancestry and ancestry-specific GWAS meta-analyses were uploaded to the Functional Mapping and Annotation (FUMA)^26^ SNP2GENE module at https://fuma.ctglab.nl/snp2gene. Except for the East Asian analysis, as it had only one contributing dataset, we restricted each analysis to variants with multiple contributing datasets (HetDf>0). MAGMA was enabled, and genomic risk loci were identified using an r^2^ > 0.1 LD threshold. FUMA-mapped genes were forwarded to the GENE2FUNC module for gene set and pathway enrichment analyses, enabling all background genes.

### LDSC

We used Linkage Disequilibrium Score Regression^79^ software to examine inflation and estimate heritability. The intercepts are reported in Results and in **Supplementary Table 5**. The λ_GC_ values from LDSC were 1.07 for multi-, 1.05 for European, 1.00 for East Asian, and 1.07 for African ancestry analyses. We also used LDSC to estimate SNP-based heritability for each ancestry-specific meta-analysis, using corresponding GWAS summary statistics. We estimated on the liability scale, using previously published approximate population prevalences^80^ to conduct our analyses.

### Genome-wide Complex Trait Analysis (GCTA)

Joint and conditional analysis was performed with GCTA.^81^ We utilized the -cojo method and set the significance threshold at 5.0×10^-8^, performing analyses per chromosome and combining results to form the set of jointly independent significant signals. In the multi-ancestry analysis, SNPs represented by only one dataset (except for the East Asian analysis, which consists only of summary statistics from Biobank Japan) were excluded to attain a conservative estimate of independent loci with multiple lines of supporting evidence. Loci were considered conditionally independent loci if: p<5.0×10^-8^ in both the meta-analysis (p) and the conditional (p_cond) analysis; -log10(p) / -log10(p_cond) < 1.5, less than a 1.5-fold difference between meta- and conditional analyses; and the lead SNP was not in LD (r^2^ > 0.1) with any other lead SNPs.^82^

### Genetically Predicted Gene Expression (GPGE)

We investigated the gene expression effects of keloid-associated variants with S-PrediXcan^34,83^ using 49 tissues from GTEx v8.^35^ The threshold for statistical significance (all tissues) was defined as 1.8×10^-7^, determined using the number of gene models and tissues analyzed. Results were further filtered to examine associations with predicted expression in particular tissues of interest, including sun-exposed skin from the lower leg, not sun-exposed skin from the suprapubic region, cell-cultured fibroblasts, and whole blood. The significance threshold for the four keloid-relevant tissues was p<1.5×10^-6^. We additionally performed colocalization analyses to test the hypothesis that a single variant is responsible for both the GWAS signal and the predicted expression association identified in GPGE. Coloc,^84^ a Bayesian gene-level test, was used to compare the GWAS and GPGE summary statistics. A statistically significant GPGE result plus a posterior probability of 90% (PP>0.90) or greater was considered strong evidence of colocalization.

### IPA

GPGE summary statistics were filtered to results achieving nominal significance (p<0.05), then analyzed using the core analysis function in Ingenuity Pathway Analysis (IPA) software (Qiagen).^36^ IPA was run for the multi-ancestry and ancestry-specific analyses, with results for networks, pathways, and upstream regulators ordered by enrichment p-value.

## Acknowledgments

CAG was supported by TL1TR002244.

GH was supported by T32GM080178.

MMS and JNH were supported by K12AR084232 (Principal Investigator: DRVE).

DRVE was supported by R21AR067938.

The eMERGE Network is supported by numerous grants from the NHGRI including U01HG004438 (CIDR) and U01HG004424 (the Broad Institute). BioVU is supported by institutional funding, the 1S10RR025141-01 instrumentation award, and by the CTSA grant UL1TR000445.

The authors acknowledge the UK Biobank, FinnGen, Biobank Japan, and the Million Veteran Program for graciously providing their GWAS summary statistics to the research community. The authors additionally thank MVP staff, researchers, volunteers, and especially participants who previously served their country in the military and now generously agreed to enroll in the study. (See https://www.research.va.gov/mvp/ for more details). Thanks to all who contributed to this work, as well as to those who made their summary statistics publicly available.

## AUTHOR CONTRIBUTIONS STATEMENT

**Conceptualization**: CAG, TLE, DRVE, JNH; **Data curation**: CAG, TLE, DRVE, JNH; **Formal analysis**: CAG, JJ; **Funding acquisition**: TLE, DRVE; **Investigation**: CAG, GH, MMS; **Project administration**: DRVE, JNH; **Resources**: AK, YL, GPJ, BNK; **Supervision**: TLE, DRVE, JNH; **Visualization**: CAG; **Writing – original draft**: CAG, TLE, DRVE, JNH; **Writing – review and editing**: CAG, GH, JJ, MMS, AK, YL, GPJ, BNK, TLE, DRVE, JNH.

## DATA AVAILABILITY

Summary statistics for the multi-ancestry and ancestry-specific meta-analyses will be deposited in the GWAS Catalog. Study-specific summary statistics for BioBank Japan [https://www.pheweb.jp/pheno/Keloid] and FinnGen [https://r8.finngen.fi/pheno/L12_HYPETROPHICSCAR] are available at their respective web portals. Summary statistics for MVP are available under dbGaP study accession phs002453.v1.p1 [https://ftp.ncbi.nlm.nih.gov/dbgap/studies/phs002453/analyses/HARE/]. UKBB data access can be requested [https://www.ukbiobank.ac.uk/enable-your-research/apply-for-access]. BioVU [https://victr.vumc.org/how-to-use-biovu/] and eMERGE [https://emerge-network.org/collaborate/] data also require approved access, which can be requested at their respective links. The predicted expression models [https://predictdb.org/post/2021/07/21/gtex-v8-models-on-eqtl-and-sqtl/] used are publicly available. The other data generated in this study are provided in the Supplementary Data files.

---

**Supplementary Figure 1.**
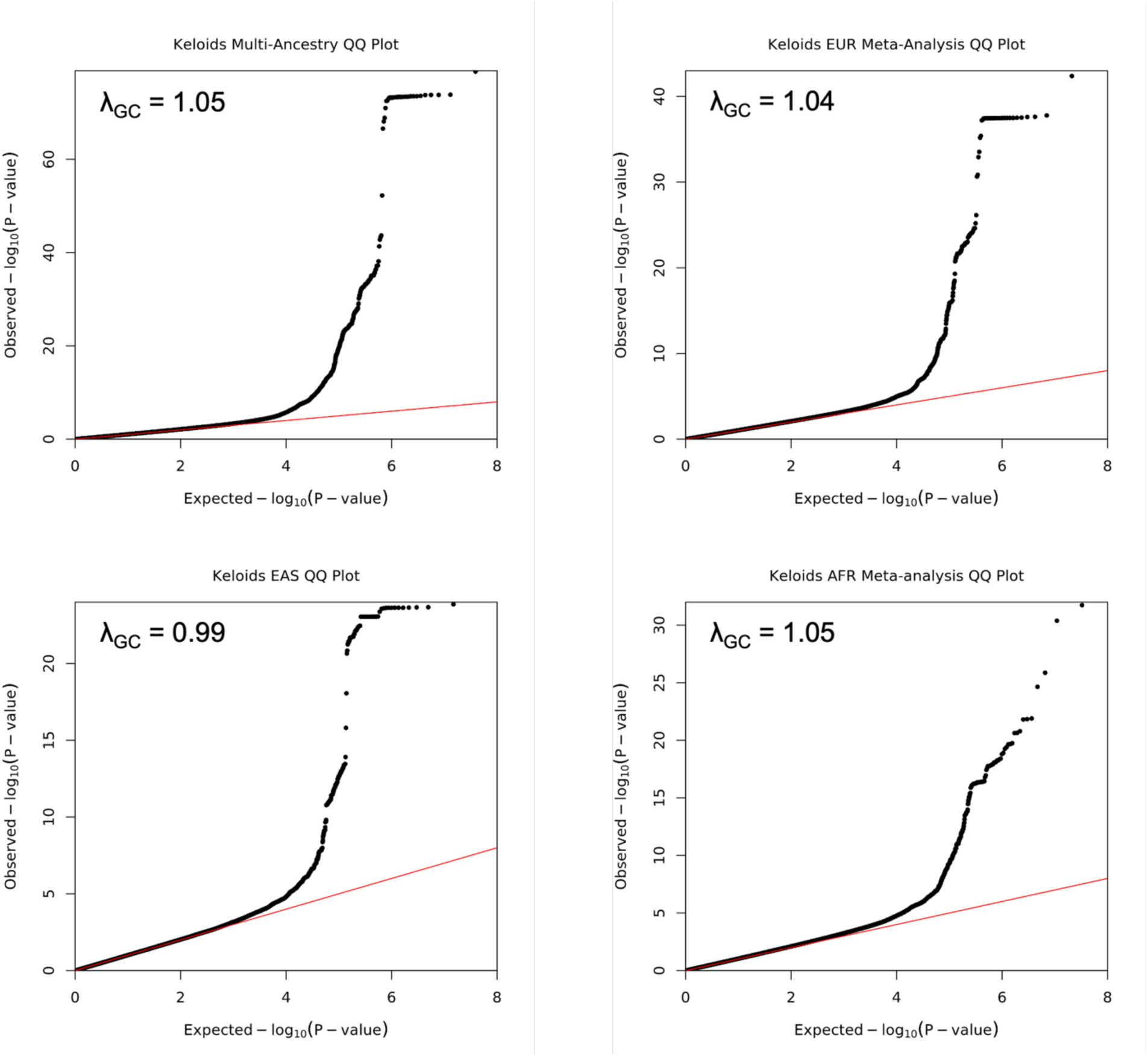
Quantile-quantile (Q-Q) plots comparing the observed distribution of p-values from each meta-analysis against an expected normal distribution. Plots created (including λ_GC_ values) using fastman R library.

**Supplementary Table 2.**
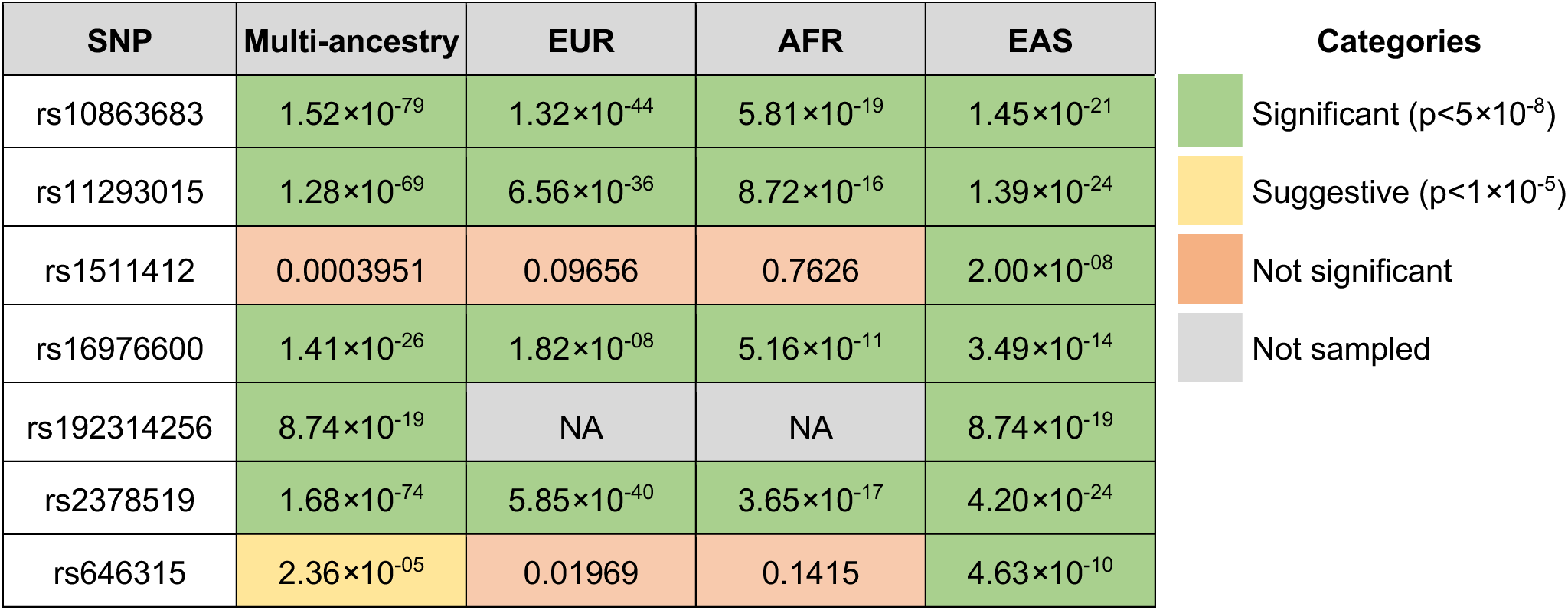
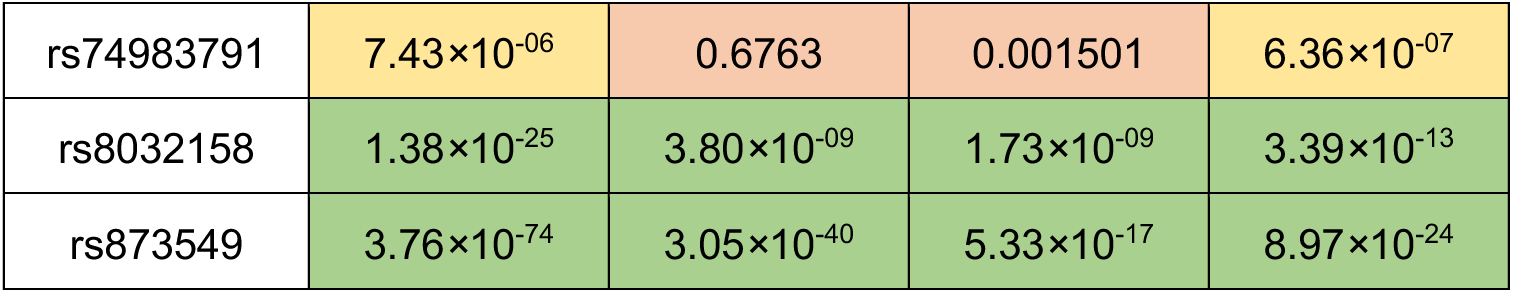
Replication status of previously identified genome-wide significant (p<5×10^-8^) SNPs. Of the 10 previously identified variants, seven were replicated at genome-wide significance in the cross-ancestry analysis. The remaining three variants had suggestive evidence of association.

**Supplementary Table 4.** Lead SNPs for each of the ancestry-specific analyses (European=EUR; East Asian=EAS; African=AFR). All alleles and effect sizes are reported in the risk-increasing direction, except for the EAS indel rs11293015.

| Ancestry | SNP | Chr:BP | Effect | Ref | OR (95% CI) | P-value | Frequency |  |
| --- | --- | --- | --- | --- | --- | --- | --- | --- |
|  |  |  |  |  |  |  | GWAS | gnomAD |
| EUR | rs35383942 | 1:201437832 | T | C | 1.49 (1.38 — 1.61) | 1.11×10<sup>-23</sup> | 0.0862 | 0.1185 |
| EUR | rs10863683 | 1:222251089 | C | G | 1.39 (1.33 — 1.46) | 1.32×10<sup>-44</sup> | 0.3310 | 0.3236 |
| EUR | rs4980386 | 11:1895708 | C | A | 1.15 (1.10 — 1.20) | 1.12×10<sup>-09</sup> | 0.6081 | 0.6126 |
| EUR | rs8032158 | 15:56194877 | C | T | 1.15 (1.10 — 1.20) | 3.80×10<sup>-09</sup> | 0.3122 | 0.3180 |
| EUR | rs34647667 | 15:68789866 | T | G | 1.30 (1.21 — 1.39) | 6.18×10<sup>-14</sup> | 0.8569 | 0.8482 |
| EUR | rs72905568 | 18:42245105 | C | T | 1.30 (1.19 — 1.41) | 2.15×10<sup>-09</sup> | 0.9154 | 0.9190 |
| EUR | rs56238684 | 20:33236696 | C | G | 1.33 (1.22 — 1.44) | 7.87×10<sup>-11</sup> | 0.0733 | 0.0733 |
| EAS | rs192314256 | 1:201437730 | C | T | 6.21 (4.14 — 9.30) | 8.74×10<sup>-19</sup> | 0.0152 | 0.0133 |
| EAS | rs10863683 | 1:222251089 | C | G | 1.61 (1.46 — 1.78) | 1.45×10<sup>-21</sup> | 0.3187 | 0.3542 |
| EAS | rs11293015 | 1:222267442 | G | GT | 0.60 (0.54 — 0.66) | 1.39×10<sup>-24</sup> | 0.7133 | 0.6499 |
| EAS | rs374569112 | 1:223403377 | G | GC | 2.42 (1.62 — 3.63) | 1.85×10<sup>-05</sup> | 0.0136 | 0.0083 |
| EAS | rs646315 | 3:138842704 | T | G | 1.75 (1.47 — 2.09) | 4.63×10<sup>-10</sup> | 0.0707 | 0.0349 |
| EAS | rs16976600 | 15:56154635 | T | C | 1.43 (1.30 — 1.56) | 3.49×10<sup>-14</sup> | 0.3564 | 0.3058 |
| AFR | rs10863683 | 1:222251089 | C | G | 1.34 (1.26 — 1.43) | 5.81×10<sup>-19</sup> | 0.2753 | 0.2506 |
| AFR | rs191467669 | 1:222501109 | A | G | 1.79 (1.50 — 2.14) | 1.01×10<sup>-10</sup> | 0.0202 | 0.0113 |
| AFR | rs76024540 | 11:2920108 | T | C | 1.47 (1.35 — 1.61) | 2.54×10<sup>-18</sup> | 0.1085 | 0.1188 |
| AFR | rs59382910 | 15:56273565 | CT | C | 1.27 (1.19 — 1.35) | 1.13×10<sup>-12</sup> | 0.3372 | 0.3545 |
| AFR | rs34647667 | 15:68789866 | T | G | 1.48 (1.39 — 1.58) | 1.83×10<sup>-32</sup> | 0.6576 | 0.6481 |
| AFR | rs140716753 | 20:11106003 | A | C | 1.72 (1.50 — 1.98) | 2.35×10<sup>-14</sup> | 0.0333 | 0.0356 |
| AFR | rs10854317 | 21:29817983 | A | G | 1.39 (1.25 — 1.55) | 3.78×10<sup>-09</sup> | 0.8815 | 0.8825 |

**Supplementary Figure 2.**
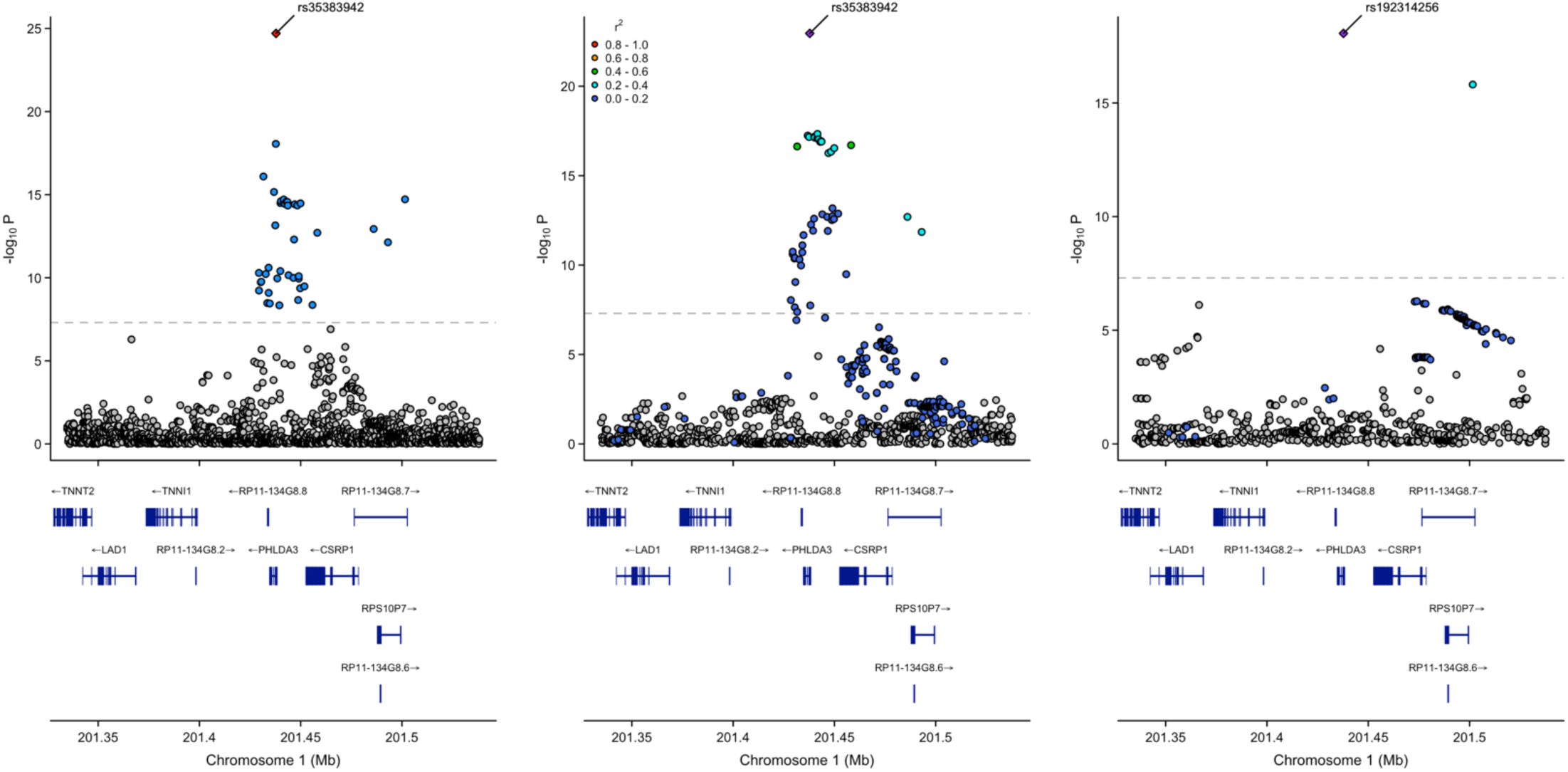
Independent genome-wide significant SNPs mapping to *PHLDA3* on chr1. Ancestry-specific analyses (**b** and **c**) also display population-specific linkage disequilibrium information. The variant rs192314256 in the East Asian analysis is relatively rare, at just above 1% in the East Asian population; The variant rs35383942 in the multi-ancestry and European ancestry analyses, meanwhile, is vanishingly rare (<0.01%) in East Asian populations.

**Supplementary Figure 3.**
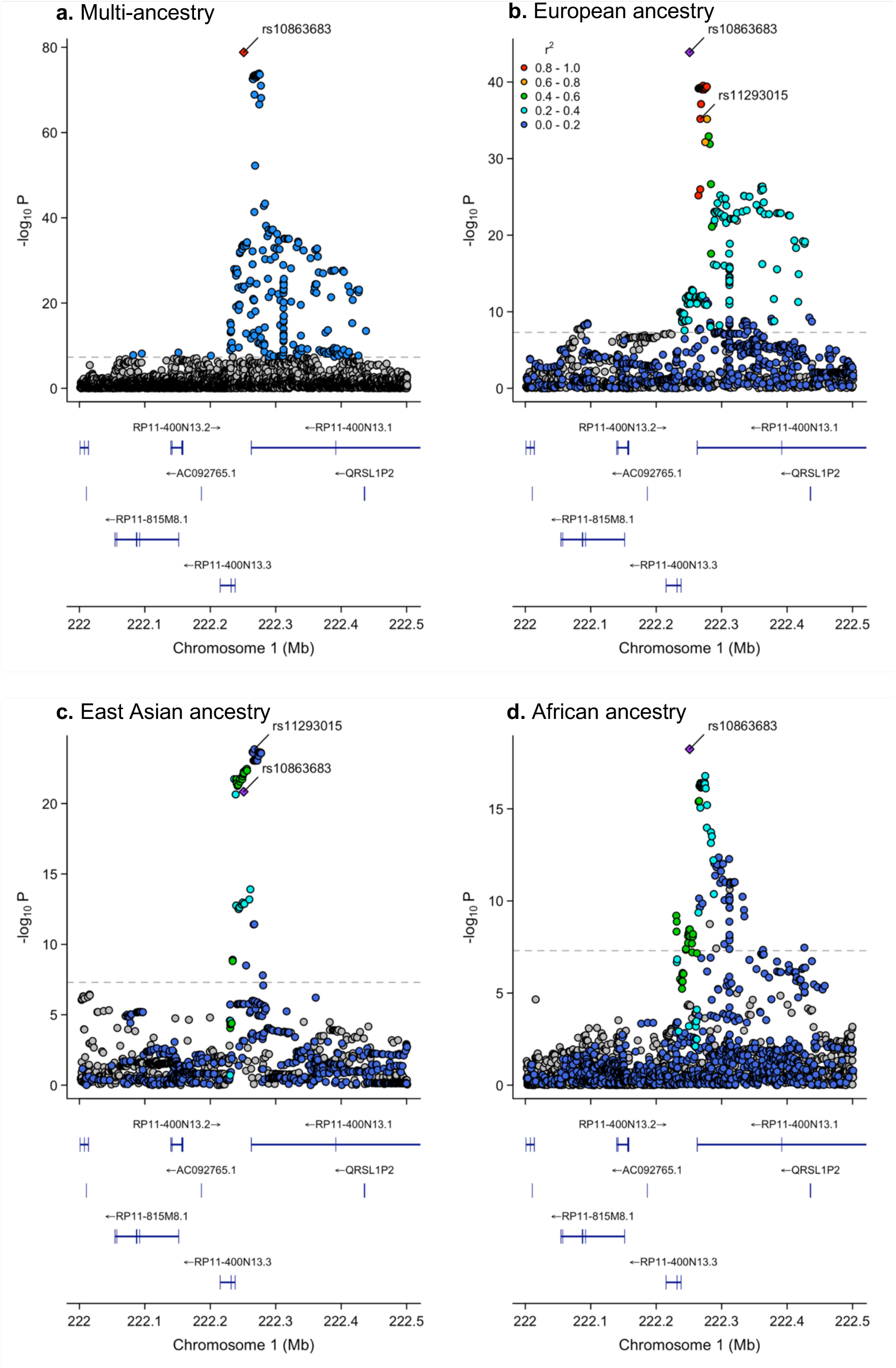
Independent genome-wide significant SNPs mapping to *RP11-415K20.1 / LINC01705* on chr1. Across all analyses, rs10863683 is a conditionally independent lead SNP. **b.** Variant rs10863683 is in linkage disequilibrium with rs11293015 in European populations and does not constitute a separate genomic risk locus in Europeans. **c.** The most significant SNP in the East Asian analysis was rs11293015, shown to be in linkage equilibrium with rs10863683 in East Asian populations. **d.** The variant rs191467669 was conditionally independent from rs10863683 in the African ancestry analysis. However, it is not present in the 1000 Genomes LD reference and is not shown here.

**Supplementary Figure 4.**
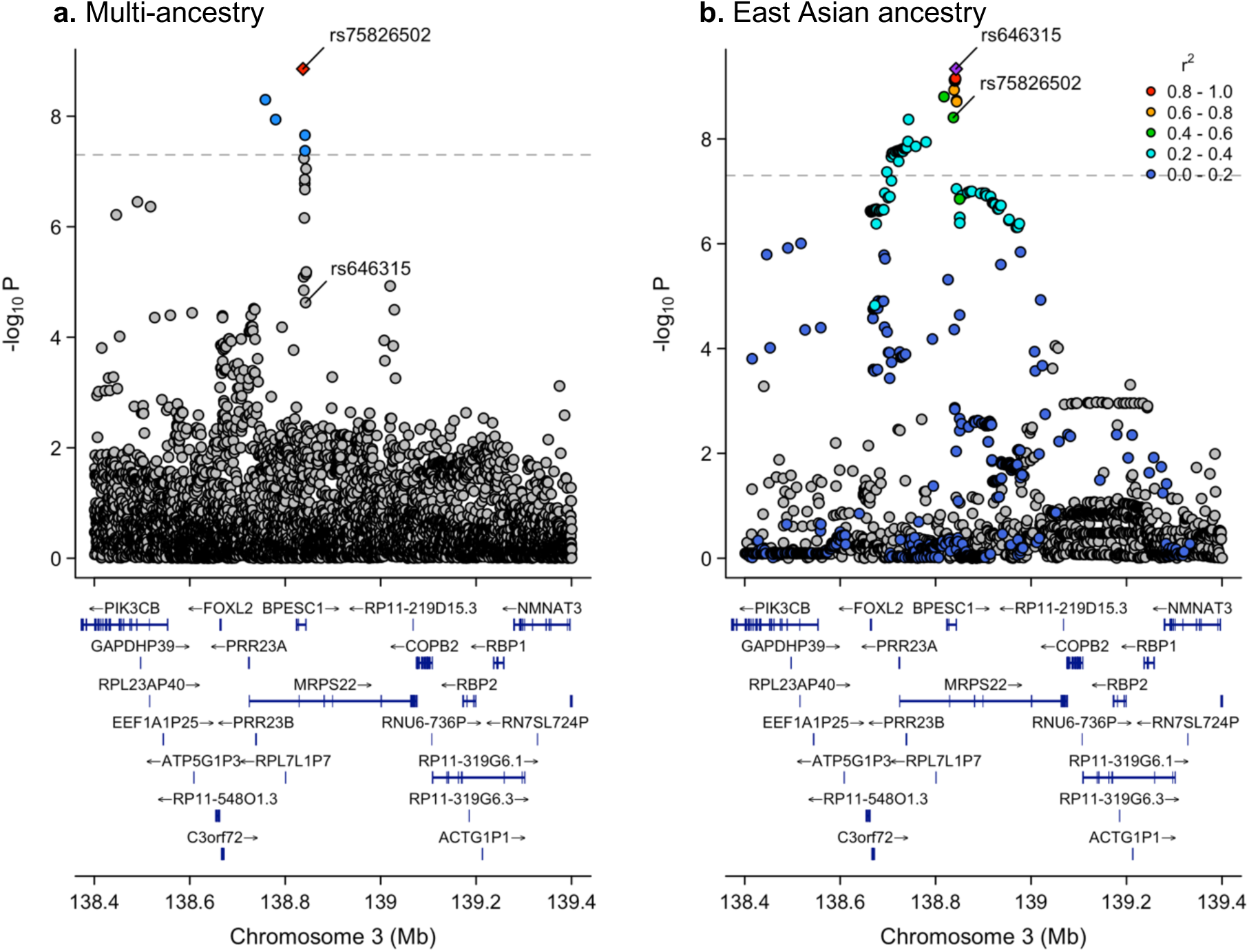
Independent genome-wide significant SNPs mapping to *MRPS22 / BPESC1* on chr3. Variant rs75826502 is the index SNP for this locus in the multi-ancestry analysis (a.), as the most significant SNP in the East Asian analysis (rs646315, **b**) was not well-supported by other datasets. This disparity does not appear to be due to allele frequency limitations in non-Asian populations, however – East Asians have the lowest population frequency of the effect allele, T (∼3%).

**Supplementary Figure 5.**
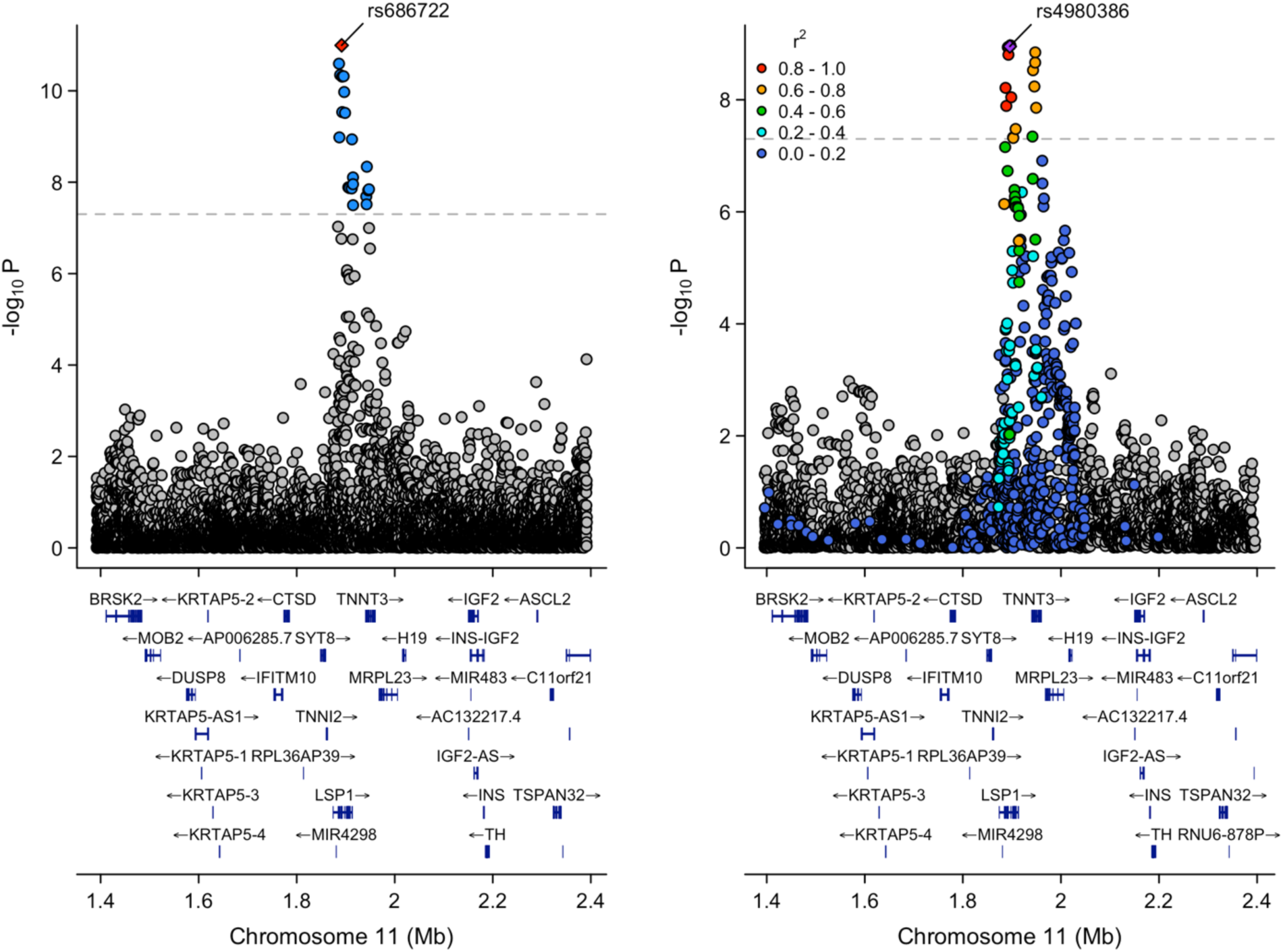
Independent genome-wide significant SNPs mapping to *LSP1* on chr11. There are different lead SNPs for the multi-ancestry (**a**) versus European ancestry analyses (**b**), which are at similar positions (approximately 4 kb apart).

**Supplementary Figure 6.**
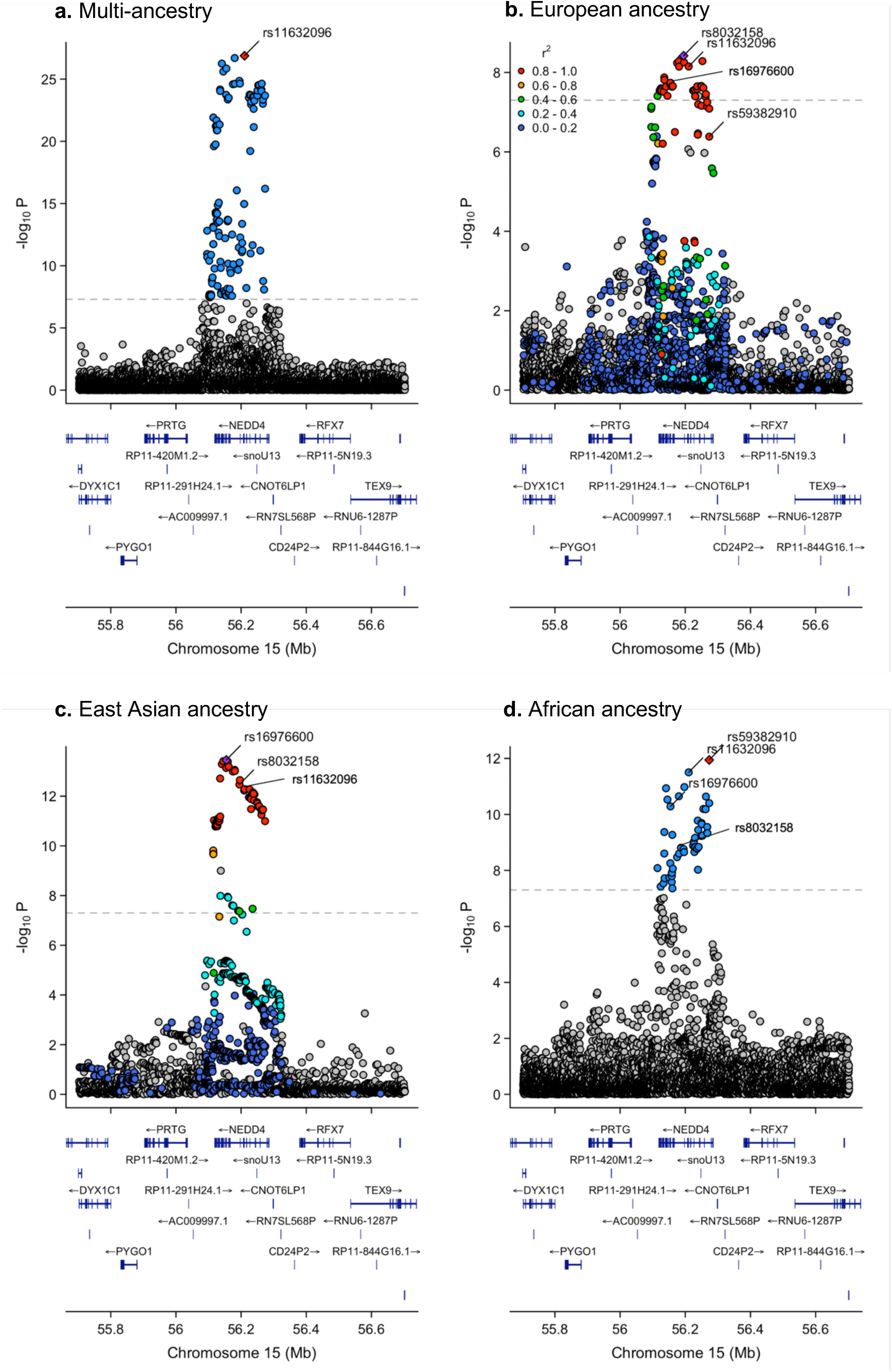
Independent genome-wide significant SNPs mapping to *NEDD4* on chr15. Each analysis had a different lead SNP representing the genomic risk locus at *NEDD4*, though they are largely in high LD with each other. All lead SNPs for each of the ancestry-specific analyses (**b-d**) are labeled, if available. The exception is the African ancestry analysis, which has other SNPs in low LD with rs59382910.

**Supplementary Table 8.**
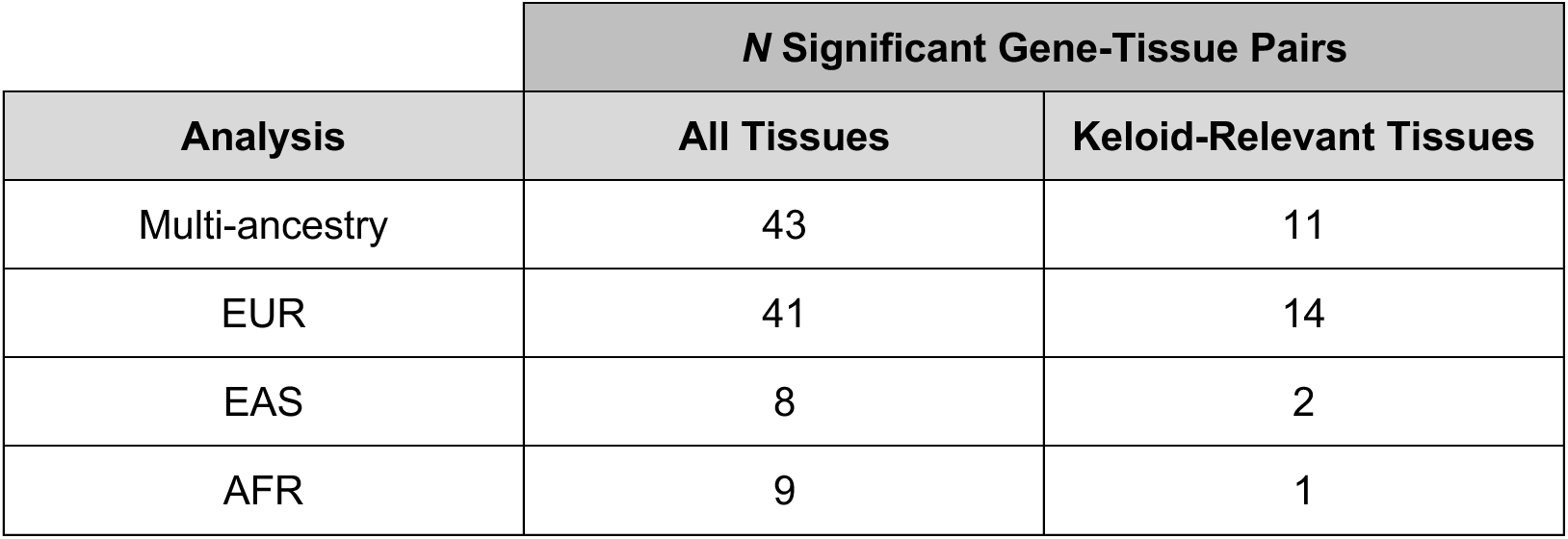
Summary of significant findings from GPGE analysis. Significance threshold (calculated based on number of tested gene-tissue pairs) for all tissues = p<1.7×10^-7^; significance threshold for the five keloid-relevant tissues = p<1.5×10^-6^.

|  | **N Significant Gene-Tissue Pairs** |  |
| --- | --- | --- |
| **Analysis** | **All Tissues** | **Keloid-Relevant Tissues** |
| Multi-ancestry | 43 | 11 |
| EUR | 41 | 14 |
| EAS | 8 | 2 |
| AFR | 9 | 1 |

